# The French Early Breast Cancer Cohort (FRESH): a resource for breast cancer research and evaluations of oncology practices based on the French National Healthcare System Database (SNDS)

**DOI:** 10.1101/2022.03.14.22272286

**Authors:** Elise Dumas, Lucie Laot, Florence Coussy, Beatriz Grandal Rejo, Eric Daoud, Enora Laas, Amyn Kassara, Alena Majdling, Rayan Kabirian, Floriane Jochum, Paul Gougis, Sophie Michel, Sophie Houzard, Christine Le Bihan-Benjamin, Philippe-Jean Bousquet, Judicaël Hotton, Chloé-Agathe Azencott, Fabien Reyal, Anne-Sophie Hamy

## Abstract

**Background:** Breast cancer (BC) is the most frequent cancer and the leading cause of cancer-related death in women. The French National Cancer Institute has created a national cancer cohort to promote cancer research and improve our understanding of cancer using the National Health Data System (SNDS). This cohort amalgamates all cancer sites, with no detailed separate data for early BC.

**Objectives:** We describe the French Early Breast Cancer Cohort (FRESH).

**Methods:** All French women aged 18 years or over, with early-stage BC newly diagnosed between January 1, 2011 and December 31, 2017, treated by surgery and registered in the general health insurance coverage plan were included in the cohort. Patients with suspected locoregional or distant metastases at diagnosis were excluded. BC treatments (surgery, chemotherapy, targeted therapy, radiotherapy, endocrine therapy), and diagnostic procedures (biopsy, cytology, imaging) were extracted from hospital discharge reports, outpatient care notes or pharmacy drug delivery data. BC subtype was inferred from the treatments received.

**Results:** We included 235,368 patients with early BC in the cohort (median age: 60 years). BC subtype distribution was as follows: luminal (80.2%), triple-negative (TNBC, 9.5%); *HER2*^+^ (10.3%), or unidentifiable (*n*=44,388, 18.9% of the cohort). Most patients underwent radiotherapy (*n*=200,685, 85.3%) and endocrine therapy (*n*=165,655, 70.4%), and 38.3% (*n*=90,252) received chemotherapy. Treatments and care pathways are described.

**Conclusion:** The FRESH Cohort is an unprecedented population-based resource facilitating future large-scale real-life studies aiming to improve care pathways and quality of care for BC patients.

## Introduction

Breast cancer (BC) is the most frequent cancer in French women, and the leading cause of cancer-related death in women. BC is treated by surgery, radiotherapy, chemotherapy and endocrine therapy, which, together with the targeted therapies developed in recent decades, have greatly improved overall survival. Following the recent FDA approval of pembrolizumab, in combination with chemotherapy in the neoadjuvant setting (Schmid et al., 2020), immunotherapies may be added to the therapeutic arsenal against BC in the near future.

The French national health insurance system covered 98.8% of the 67 million inhabitants of France in 2020 (Casarotto et al., 2021). All the medical and administrative information relating to the reimbursement of French citizens for healthcare expenses are collected and aggregated in the National Health Data System (*Système National des Données de Santé*, SNDS) database (Tuppin et al., 2017). In 2014, the French National Cancer Institute (Institut National du Cancer, INCa) set up the French cancer cohort, an exhaustive population-based cohort, based on SNDS data (Bousquet et al., 2018). This resource aims to provide a robust and validated database for the analysis of cancer complications, adverse effects, morbidity and mortality, and it provides opportunities for studying expenditure indicators and the monitoring of care consumption, quality and safety indicators, oncologic outcomes, geographic distributions and care pathways. This resource currently contains amalgamated data for all sites of cancer, with no detailed data available separately for early BC.

We describe here a specific subset of the French cancer cohort comprising all women diagnosed with early breast cancer, in the context of the French Early Breast Cancer Cohort (FRESH). We describe data sources, inclusion and exclusion criteria, basic descriptive analyses, and longitudinal trends over time. We performed quality control and benchmarking against published data, and we discuss perspectives for improving oncology practices, generating research hypotheses, and overcoming existing challenges.

## Materials and methods

### Data source and available variables

Relevant data were identified with the Oncology Data Platform (ODP) available at the French National Cancer Institute (INCa). The ODP gathers together SNDS data for all individuals living in France with universal health insurance cover (98.8% of the population) (Moulis et al., 2015a) who were diagnosed with or treated for cancer between 2010 and 2018. The ODP has been described in detail elsewhere (Bousquet et al., 2018). It includes (i) demographic data (sex, date of birth, zip code of the town of residence, vital status, date of death if appropriate, health insurance regimen), (ii) hospital discharge reports (diagnoses, medical procedures, and expensive treatments), (iii) outpatient care (drugs dispensed, with the date of delivery, laboratory tests, and outpatient medical procedures) from the year preceding the date of the inclusion of the patient in the ODP up to December 31, 2018 and (iv) all long-term illness (LTI) records (diagnosis codes and date of disease onset) until December 31, 2018. The Diagnosis codes were recorded with the International Classification of Diseases – 10^th^ revision, ICD-10 (World Health Organization). Procedures were recorded with the CCAM classification (*Classification Communes des Actes Médicaux*). Molecules in outpatient care were fully identifiable and were recorded with CIP (*Code Identifiant de Présentation*) codes. In hospital, only costly innovative drugs part of a special reimbursement process called “list en sus” were recorded, under the form of UCD (*Unités Communes de Dispensation*) codes. Both the UCD and CIP codes were linked to the ATC classification (Anatomical Therapeutic and Chemical classification) of the World Health Organization. In France, several health insurance coverage plans exist, depending on occupational status. The general health insurance plan (“Régime Général”) gathers approximately 88% of the French population: employees in the industry, business, and service sectors; public service employees; and students.

### Ethics and data protection

This study was conducted in the framework of a partnership between Institut Curie and the French INCa. It was performed in accordance with institutional and ethical rules concerning research using data from patients. The study was authorized by the French data protection agency (*Commission nationale de l’informatique et des libertés*—CNIL, under registration number 920017). Personal data were kept confidential, for all participants, by pseudoanonymizing the data. Written informed consent from the patients was not required, in accordance with French regulations.

### Selection of the patients

We applied 10 filters for patient inclusion in the cohort (Fig. S1). We identified patients with (1) BC (2) newly diagnosed between January 1, 2011 and December 31, 2017. We also applied sociodemographic filters to exclude: (3) male patients, as they represent a very specific population with different therapeutic approaches (Gucalp et al., 2019)), (4) patients under the age of 18 years at inclusion, (5) patients who were not registered in the general health insurance coverage plan (ensuring exhaustivity for vital status outcomes). Patients who did not undergo breast surgery in the year following inclusion (6) were excluded from the cohort to ensure the exclusion of relapses of a previous BC diagnosis. Such exclusion criteria was unlikely to bias significantly the population cohort, as more than 95% of patients treated for an incident non-metastatic BC in France received surgery ((Bihan-Benjamin et al., 2021)). It also enabled fixing an index surgery date for use as a reference for the definition and the settings (neoadjuvant *versus* adjuvant) of the other BC treatments. We excluded patients with other concomitant cancers (7) to ensure that the retrieved cancer treatments are BC treatments. Patients with evidence of a previous BC (8) or evidence of stage IV metastatic BC at diagnosis (9) were also excluded from the cohort (metastatic BC is an incurable disease with specific chemotherapy and targeted therapy molecules, divergences in medical opinions regarding need for surgery (Tosello et al., 2018) and a low median survival of about 3 years (Gobbini et al., 2018)). Finally, we excluded patients with poor-quality or inconsistent data (10). The full process of patient selection is detailed in the Supplemental Material and in Tables S1-3.

### BC treatments

#### Surgery

Surgery for BC was tagged with CCAM procedure codes (Table S4) at the hospital, with classification into five categories: (1) mastectomy with axillary surgery, (2) mastectomy without axillary surgery, (3) partial mastectomy with axillary surgery, (4) partial mastectomy without axillary surgery and (5) axillary surgery without breast surgery. The *index surgery* for BC was defined as the date on which the first breast surgical operation for BC (categories 1 to 4 above) took place, in the year of inclusion or the following year. This date was used as a reference for the definition of the other treatments. The decision rules used to bin breast surgery types (partial mastectomy *versus* mastectomy) and axillary surgery (yes *versus* no) are detailed in the Supplemental Material, together with the rules for data handling for patients undergoing several surgical procedures.

#### Radiotherapy

*Radiotherapy (RT) sessions* were identified by ICD-10 diagnosis codes or CCAM procedure codes (TableS4). We used temporal restrictions to exclude RT sessions for another cancer or a BC relapse: a patient was considered to have been treated with RT if she had at least one RT session between 150 days before and up to 365 days after BC index surgery. Thresholds have been set in accordance with clinical practices. Decision rules concerning the *radiotherapy setting (neoadjuvant / adjuvant / both)* and the identification of start and end dates are detailed in the Supplemental Material.

#### Chemotherapy

*Chemotherapy (CT) sessions* were identified by ICD-10 diagnosis codes, CCAM procedure codes and ATC molecule codes for hospital and outpatient care (Table S4). A patient was considered to have been treated with CT if she had at least one CT session between 250 days before and up to 180 days after BC index surgery, in accordance with clinical practices. Decision rules for *chemotherapy setting (neoadjuvant / adjuvant / both)* and the identification of start and end dates, and intervals between CT sessions are detailed in the Supplemental Material.

The following seven *CT regimens* were identified: (1) anthracyclines, (2) anthracyclines/docetaxel, (3) anthracyclines/paclitaxel, (4) docetaxel, (5) paclitaxel, (6) unknown or (7) other; as detailed in the Supplemental Material, together with the number of cycles, and displayed in Fig. S2. Regimens containing paclitaxel were fully identifiable, whereas regimens containing only anthracyclines and/or docetaxel were not and were mostly tagged as “Unknown”.

#### Endocrine therapy

*Endocrine therapy (ET*) *intake* was tagged on the basis of the outpatient delivery of (1) tamoxifen, (2) aromatase inhibitors (AI) or (3) gonadotropin-releasing hormone agonists (GnRH agonists), identified with ATC codes (Table S4). Decision rules for *endocrine therapy setting (neoadjuvant followed by adjuvant / adjuvant)*, for the exclusion of treatments linked to fertility preservation procedures, and for the identification of start and end dates are detailed in the Supplemental Material.

*ET regimens* were classified into seven categories according to the ET molecules delivered over the whole study period: (1) tamoxifen only (tamoxifen); (2) at least one delivery of tamoxifen plus at least one delivery of GnRH agonists (tamoxifen with GnRH agonists); (3) at least one delivery of tamoxifen followed by at least one delivery of AI (tamoxifen followed by AI); (4) AI only (AI); (5) at least one delivery of AI and at least one delivery of GnRH agonists (AI with GnRH agonists); (6) at least one delivery of AI followed by at least one delivery of tamoxifen (AI followed by tamoxifen); (7) all other cases, including the delivery of GnRH agonists alone, the delivery of three types of ET, or multiple sequential combinations of AI and tamoxifen (others).

#### Targeted therapy (TT)

Only two targeted therapies (TT) (trastuzumab and pertuzumab) have been approved for the early BC setting. *Anti-HER2 (human epidermal growth factor receptor 2) targeted therapy (TT) sessions* were identified by ATC codes (Table S4) for trastuzumab and/or pertuzumab. Patients were considered to have received targeted therapy if they had at least one anti-*HER2* targeted therapy session between 250 days before and up to 180 days after BC index surgery, in accordance with clinical practices. The decision rules for *TT setting (neoadjuvant followed by adjuvant / adjuvant)* and the identification of start and end dates are detailed in the Supplemental Material. TT regimens were classified as: (1) trastuzumab alone (trastuzumab) or (2) pertuzumab with or without trastuzumab (pertuzumab +/-trastuzumab). The decision rules for combinations of TT with other systemic treatments and the number of cycles are detailed in the Supplemental Material and displayed in Fig. S3. Of note, the diagnostic code of chemotherapy sessions does not enable distinguishing sessions of TT combined with chemotherapy and sessions of TT alone.

*Combinations of TT and systemic treatments* were classified as follows: (1) Anthracycline-based regimen alone and then a combination of docetaxel and TT (anthracyclines/docetaxel-TT); (2) anthracycline-based regimen alone and then a combination of paclitaxel and TT (anthracyclines/paclitaxel-TT); (3) combination of a docetaxel-based regimen and TT (docetaxel-TT); (4) combination of paclitaxel and TT, as described by Tolaney et al. (Tolaney et al., 2015), (paclitaxel-TT (Tolaney)); (5) combination of endocrine therapy and TT, without chemotherapy (TT-ET); (6) any other combinations including TT (other).

### Date of first BC treatment

We defined the date of first BC treatment as the date of BC index surgery or the start date of neoadjuvant chemotherapy (NAC), neoadjuvant endocrine therapy (NET), neoadjuvant radiotherapy (NRT) or neoadjuvant anti-*HER2* targeted therapy (NTT), whichever occurred first.

### BC diagnostic procedures and date of BC diagnosis

Procedures for BC diagnosis were identified by CCAM procedure codes in the hospital and outpatient settings (Table S6) and were classified into three categories: breast core biopsy, fine-needle aspiration cytology and breast imaging procedures, including mammography, mammary ultrasound, mammary MRI, CT scan and galactography. If at least one breast core biopsy had been performed in the 12 months preceding the date of first BC treatment, we considered the diagnosis of BC to be set by the earliest breast core biopsy within this time range. Otherwise, we considered it to be set by the earliest breast fine-needle cytology aspiration. If neither breast core biopsy nor fine-needle aspiration cytology had been performed, the diagnosis was assumed to have been based on a breast imaging procedure. In such cases, the BC diagnosis date was set as the date of the earliest of such procedures, starting from the breast imaging procedure closest to the date of first BC treatment and going back breast imaging procedures one by one, as long as they are separated by no more than a month interval. The date of first BC treatment was taken as the date of diagnosis if no diagnostic procedure was recorded.

#### Variables of interest

##### Sociodemographic variables

*Age at diagnosis* was calculated by the rounded difference, in years, between the date of BC diagnosis and the date of birth.

##### Inferred BC subtype

No information about the histological characteristics of the tumor was available from the database. We therefore inferred BC subtype from the treatments received. The tumors of patients receiving anti-*HER2* TT were classified as ***HER2***^**+**^. Within this category, the tumors of patients receiving ET were classified as ***HER2***^***+***^**/HR**^**+**^, and those of patients not receiving ET were classified as ***HER2***^***+***^**/HR**^**-**^. The tumors of patients who received ET without anti-*HER2* TT were considered to be **luminal**. The tumors of patients receiving chemotherapy with no ET or anti-*HER2* TT were classified as **triple-negative breast cancers** (**TNBCs**). Finally, the tumors of patients treated exclusively by surgery with or without radiotherapy were considered to have an **undefined** subtype.

##### Nodal status

Lymph node involvement was tagged by the presence of at least one ICD-10 diagnosis code for node disease (C773) between 250 days before and up to 180 days after BC index surgery.

### Statistical analysis

Analyses were performed with R software, version 3.6.3. The study population was described in terms of frequencies for qualitative variables, or medians and interquartile ranges (IQR) for quantitative variables. For variables suspected to have a multimodal distribution based on graphical assessment, we performed a statistical test based on the critical bandwidth statistic with the *modetest* function of the R package multimode (Ameijeiras-Alonso et al., 2021). If the null hypothesis of unimodality was rejected in statistical tests, the number of modes was assessed graphically, and the position of the modes was inferred by kernel density estimation with gaussian kernels (R multimode package; function *locmodes*). The threshold for statistical significance threshold was set at *p* = 0.05. We prevented undesirable edge effects, by restricting figures with a continuous variable (age, date of diagnosis, chemotherapy start date, etc.) on the *x*-axis to strata of the continuous variable (month-years for dates, integer units for age) containing at least 50 patients.

*Overall survival* (OS) was defined as the time, in months, from BC index surgery to death or to March 1^st^, 2019, whichever occurred first. Data for patients still alive on March 1^st^, 2019 were censored at this date. Median follow-up and its interquartile range (IQR) were assessed by reverse Kaplan-Meier methods. Unadjusted survival probabilities were estimated by the Kaplan-Meier method, and survival curves were compared in log-rank tests.

## Results

### Patients and tumor characteristics

#### Age at BC diagnosis

We included 235,368 women from the 455,711 patients with a diagnosis code of BC identified in the final cohort (Fig. S1). The characteristics of the patients are summarized in Table 1. Median age at diagnosis was 60 years. (Fig. 1A). The age distribution of the patients was bimodal, with two incidence peaks, at 50.3 and 65.0 years, respectively. The value of the second mode of the distribution increased steadily over time, from 62.7 years in 2011 to 68.0 years in 2017 (Fig. S4).

**Table 1:** Baseline characteristics of the French Early Breast Cancer Cohort. Abbreviations: HR^+^ = hormone receptor-positive; HR^−^ = hormone receptor-negative; *HER2*^*+*^= human epidermal growth factor receptor 2-positive; TNBC = triple-negative breast cancer subtype; AI = aromatase inhibitor; *: chemotherapy regimens are displayed for the 91,938 (72939+15627+2*1686) chemotherapy settings identified. **: 3 of 19,722 with targeted therapy received pertuzumab only.

| Variable | Class | All |
| --- | --- | --- |
| n = |  | 235 368 |
| Age at diagnosis (years) |  | 60.0 (19.0) |
| Age at diagnosis (years, classes) | [0 - 30) | 1 124 (0.5%) |
|  | [30 - 40) | 10 539 (4.5%) |
|  | [40 - 50) | 43 206 (18.4%) |
|  | [50 - 60) | 58 003 (24.6%) |
|  | [60 - 70) | 64 042 (27.2%) |
|  | [70 - 80) | 39 163 (16.6%) |
|  | 80+ | 19 291 (8.2%) |
| Inferred BC subtype | Luminal | 153 109 (65.1%) |
|  | TNBC | 18 149 (7.7%) |
|  | HER2+ /HR+ | 12 561 (5.3%) |
|  | HER2+ /HR- | 7 161 (3.0%) |
|  | Undefined | 44 388 (18.9%) |
| Nodal status | Node negative | 191 164 (81.2%) |
|  | Node positive | 44 204 (18.8%) |
| Surgery | No | 0 (0%) |
|  | Yes | 235 368 (100%) |
|  | **Surgery type** |  |
|  | Partial Mastectomy | 173 173 (73.6%) |
|  | Mastectomy | 62 195 (26.4%) |
|  | **Axillar surgery** |  |
|  | No | 38 231 (16.2%) |
|  | Yes | 197 137 (83.8%) |
| Chemotherapy | No | 145 116 (61.7%) |
|  | Yes | 90 252 (38.3%) |
|  | **Setting** |  |
|  | Neoadjuvant | 15 627 (17.3%) |
|  | Adjuvant | 72 939 (80.8%) |
|  | Both | 1 686 (1.9%) |
|  | **Regimen\*** |  |
|  | Anthracyclines | 773 (0.8%) |
|  | Anthracyclines/Docetaxel | 13 897 (15.1%) |
|  | Anthracyclines/Paclitaxel | 21 605 (23.5%) |
|  | Docetaxel | 4 974 (5.4%) |
|  | Paclitaxel | 2 869 (3.1%) |
|  | Other | 6 441 (7.0%) |
|  | Unknown (Anthracyclines or Docetaxel) | 41 379 (45.0%) |
| Targeted therapy | No | 215 646 (91.6%) |
|  | Yes | 19 722 (8.4%) |
|  | **Setting** |  |
|  | Neoadjuvant followed by adjuvant | 4 424 (22.4%) |
|  | Adjuvant | 15 298 (77.6%) |
|  | **Regimen** |  |
|  | Trastuzumab only | 19 289 (97.8%) |
|  | Pertuzumab +/- trastuzumab** | 433 (2.2%) |
| Radiotherapy | No | 34 683 (14.7%) |
|  | Yes | 200 685 (85.3%) |
|  | **Setting** |  |
|  | Neoadjuvant | 323 (0.2%) |
|  | Adjuvant | 200 180 (99.7%) |
|  | Both | 182 (0.1%) |
| Endocrine therapy | No | 69 713 (29.6%) |
|  | Yes | 165 655 (70.4%) |
|  | **Setting** |  |
|  | Neoadjuvant followed by adjuvant | 2 438 (1.5%) |
|  | Adjuvant | 163 217 (98.6%) |
|  | **Regimen** |  |
|  | AI | 102 757 (62.0%) |
|  | Tamoxifen | 35 187 (21.2%) |
|  | Tamoxifen followed by AI | 10 350 (6.2%) |
|  | AI followed by tamoxifen | 8 011 (4.8%) |
|  | Tamoxifen in combination with agonist | 1 064 (0.6%) |
|  | AI in combination with agonist | 705 (0.4%) |
|  | Others | 7 581 (4.6%) |

**Fig. 1:**
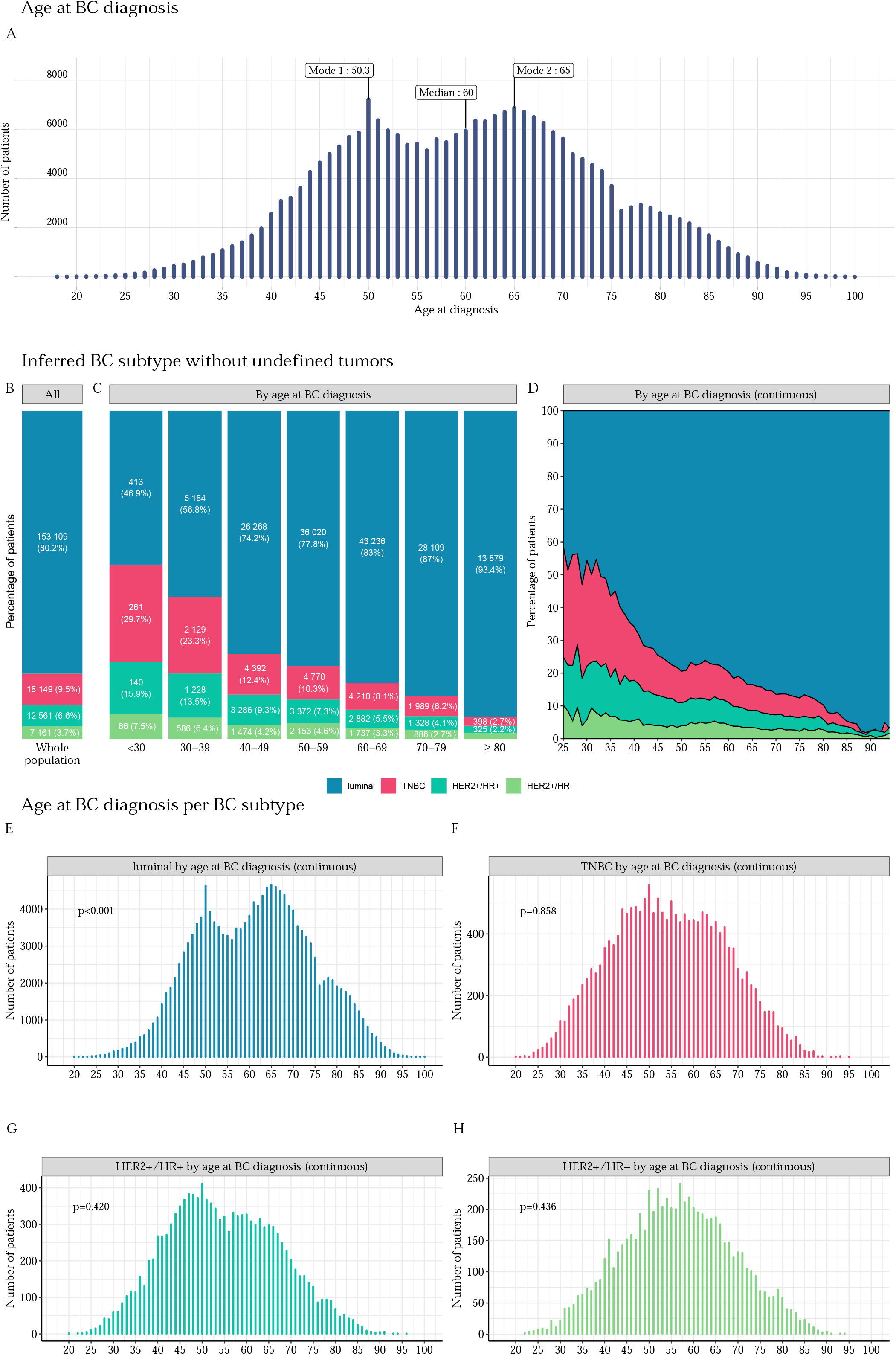
Age at BC diagnosis and inferred BC subtype, by age at BC diagnosis, excluding undefined tumors. (A) Number of patients included in the FRESH cohort, by age at BC diagnosis. The age distribution is bimodal, with two inferred incidence peaks at 50.3 and 65.0 years (*p*-value for non-unimodality < 0.001). Median age is 60 years; (B) Inferred BC subtype percentages for the whole population, excluding undefined tumors (*n*=190,980); (C) Inferred BC subtype percentages per age class at BC diagnosis, excluding undefined tumors. Raw figures for subgroups representing less than 2% of the corresponding age class are not displayed on the graph, to ensure readability. For the group ≥ 80 years old: *n*=259 (1.7%) for the *HER2*^*+*^/HR^−^ group; (D) Inferred BC subtype percentage by age at BC diagnosis, excluding undefined tumors. The cohort is restricted to patients aged from 25 to 94 years (*n*=190,816); (E) Age distribution of patients with an inferred luminal subtype tumor (*n*=153,109) at BC diagnosis (*p*-value for non-unimodality < 0.001); (F) Age distribution of patients with an inferred TNBC subtype tumor (*n*=18,149) at BC diagnosis (*p*-value for non-unimodality= 0.858); (G) Age distribution of patients with an inferred *HER2*^*+*^/HR^+^ subtype tumor (*n*=12,561) at BC diagnosis (*p*-value for non-unimodality = 0.420); (H) Age distribution of patients with an inferred *HER2*^*+*^/HR^−^ subtype tumor (*n*=7,161) at BC diagnosis (*p*-value for non-unimodality =0.436). Abbreviations: BC = breast cancer; HR^+^ = hormone receptor-positive; HR^−^ = hormone receptor-negative; TNBC = triple-negative breast cancer subtype; *HER2*^*+*^ = human epidermal growth factor receptor 2-positive.

#### Mode of diagnosis

A procedure for pathology analysis was performed before treatment in 93.3% of patients (biopsy *n*=214,874, 91.3%; cytology *n*=4,808, 2.0%) (Fig. S5). Imaging was the sole diagnostic procedure before treatment in 5.6% of patients (*n*=13,250), and 1.0% of the patients underwent no diagnostic procedures at all before treatment (*n*=2,436). This absence of diagnostic procedures was more marked at extreme ages, in the youngest and oldest women (Fig. S5B-C), and in patients whose first treatment was surgery (Fig. S5D-F) than in patients receiving neoadjuvant treatment (Fig. 4G-I).

#### Inferred BC subtypes

It was possible to infer BC subtype in 190,980 (81.1%) whereas BC subtype was undefined for 44,388 patients (18.9% of the total cohort) (Fig. S6). The distribution of inferred BC subtypes was as follows: luminal (80.2%), TNBC (9.5%), *HER2*^*+*^ (10.3%) (Fig. 1B). The proportion of TNBC and *HER2*^+^ BC decreased with advancing age (Fig. 1C-D). The bimodal distribution of BC cases by age was also observed for the luminal and undefined subtypes, but not for the TNBC, *HER2*^*+*^/HR^+^ and the *HER2*^*+*^/HR^−^ subsets (Fig. 1E-H and Fig. S7).

#### Nodal status

Lymph node involvement was present in 18.8% of the cases (*n*=44,204) (Fig. S8A) and varied with age (Fig. S8B) and inferred BC subtype (Fig. S8C). For the patients for whom a BC subtype was inferred, the proportion of node-positive tumors was lowest for luminal BCs (21.5%), and highest for HER2^+^/HR^−^ BCs (30.1%).

### BC treatments

#### Sequence of treatments and care pathways

In accordance with the inclusion criteria, 100% of the patients underwent surgery (*n*=235,368) (Fig. 2A). The distribution of other BC treatments was as follows: radiotherapy (*n*=200,685, 85.3%), endocrine therapy (*n*=165,655, 70.4%), chemotherapy (*n*=90,252, 38.3%), and targeted therapy (*n*=19,722, 8.4%).

**Fig. 2:**
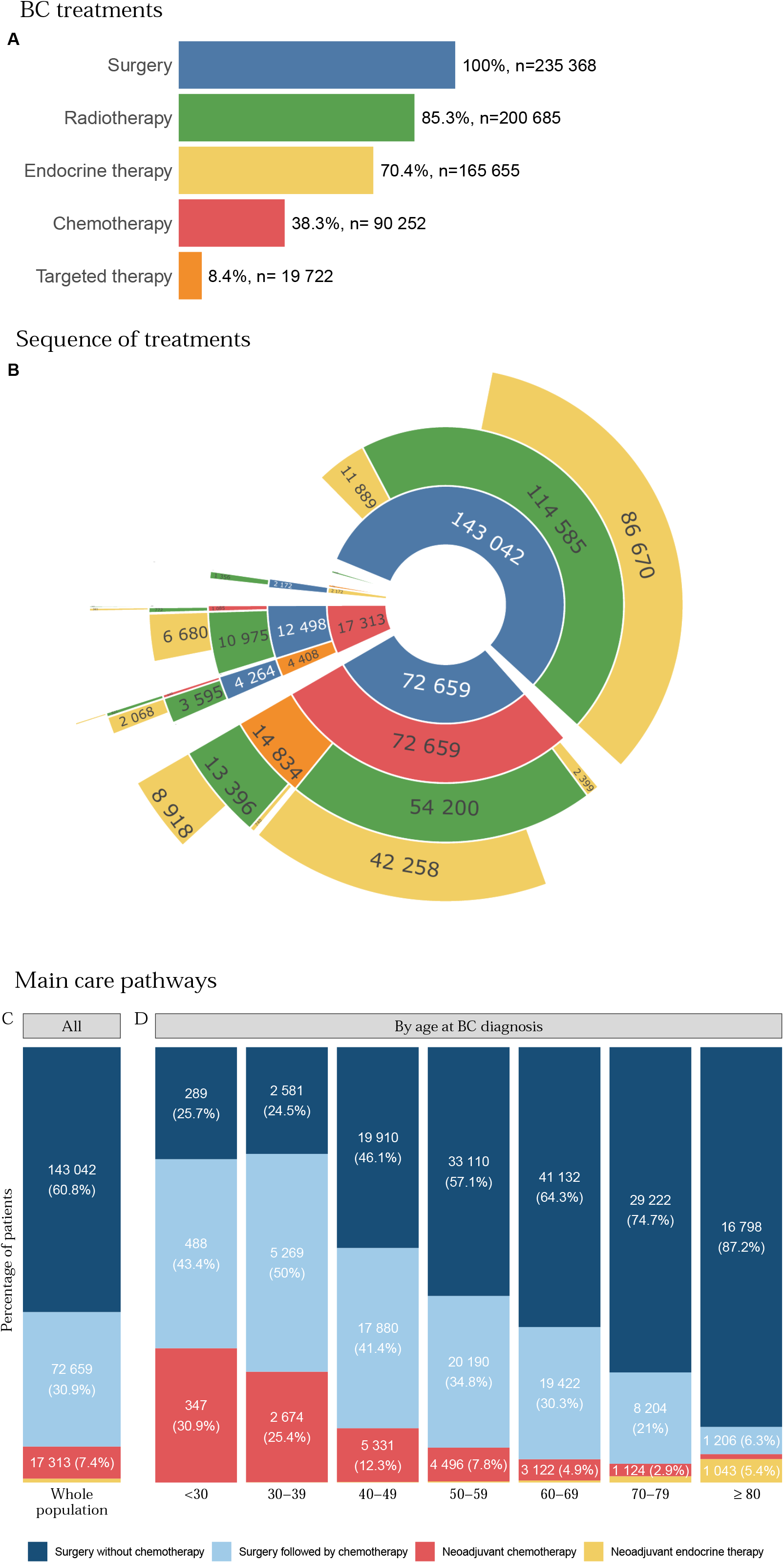
BC treatment, sequence of treatments and main care pathways, by age at BC diagnosis. (A) Distribution of BC treatment. (B) Sequential care pathways. Care pathways are displayed from inwards to outwards. For instance, n=143,042 patients received surgery first without chemotherapy. Among those, surgery was followed by radiotherapy for n=114,585 patients and by endocrine therapy without radiotherapy for n=11,889. Treatment sequences are displayed in the following order: neoadjuvant chemotherapy (NAC) - neoadjuvant targeted therapy (NTT) – neoadjuvant radiotherapy (NRT) - neoadjuvant endocrine therapy (NET) - surgery - adjuvant chemotherapy – adjuvant targeted therapy – adjuvant radiotherapy – adjuvant endocrine therapy. The continuation of NET or NTT after surgery is not considered to constitute adjuvant endocrine therapy or adjuvant targeted therapy; (C) Distribution of the four main care pathways extracted from (B): (i) surgery without chemotherapy, (ii) surgery followed by chemotherapy, (iii) neoadjuvant chemotherapy (NAC), (iv) neoadjuvant endocrine therapy (NET). For the sake of clarity, radiotherapy is not spelled out in trajectories. The vast majority of the patients in the cohort (85.3%) underwent radiotherapy as part of their care pathway. Patients with neoadjuvant chemotherapy are classified as NAC irrespective of their NRT/NET status. Patients with NRT and NET are classified as NRT (*n*=166). This category is so rare that it is not displayed on the plot. As a consequence, the patients in the NET group received neoadjuvant endocrine therapy and neither NRT nor NAC. Targeted therapy is always given in combination with either chemotherapy or endocrine therapy. This implies that NTT patients are in the NAC, NRT or NET group. Raw data for subgroups representing less than 2% of the total population are not displayed on the graph, to ensure readability: 2,169 (0.9%) patients are in the NET group; (D) Distribution of the four main treatment trajectories extracted from (B) per age class at BC diagnosis. Raw data for subgroups representing less than 2% of the corresponding age class are not displayed on the graph, to ensure readability. The values per age class are: for the <30 year-old group: *n*<10; 30-39 years old: *n*=25 (0.2%); 40-49 years old: *n*=92 (0.2%), 50-59 years old: *n*=207 (0.3%), 60-69 years old: *n*=355 (0.6%), 70-79 years old: *n*=610 (1.6%) for the NET group. For the ≥ 80 years age class: *n*=215 (1.1%) for the NAC group; Abbreviations: BC = breast cancer; CT = chemotherapy; NAC = neoadjuvant chemotherapy; NET = neoadjuvant endocrine therapy; NTT = neoadjuvant targeted therapy; NRT = neoadjuvant radiotherapy.

Four main care pathways, accounting for 99.9% of patients, were identified: (1) surgery without chemotherapy (*n*=143,042, 60.8%); (2) surgery followed by chemotherapy (*n*=72,659, 30.9%); (3) neoadjuvant chemotherapy (*n*=17,313, 7.4%); (4) neoadjuvant endocrine therapy (*n*=2,188, 0.9%) (Fig. 2B). The relative distribution of the four main care pathways is shown in Fig. 2C.

The proportion of patients treated with either neoadjuvant chemotherapy or adjuvant chemotherapy decreased with advancing age, whereas the proportion of patients treated with neoadjuvant ET increased with advancing age (Fig. 2D).

#### Locoregional treatments

##### Surgery

The distribution of surgical procedures was as follows: 73.6% of patients underwent partial mastectomy and 26.4% underwent mastectomy (Fig. S9A-D); 83.8% of patients underwent axillary surgery, and 16.2% did not (*n*=38,231) (Fig. S9E). The type of surgical procedure varied with age, BC subtype, and nodal status (Fig. S9B-D, F-H).

##### Radiotherapy

Most patients underwent radiotherapy (85.3%) (Fig. S10A), and the rate of radiotherapy varied with age, BC subtype, and nodal status (Fig. S10B-D). This rate was higher in patients treated by partial mastectomy than in patients undergoing full mastectomy (93.3% *versus* 63.0%) (Fig. S10E-H).

#### Systemic treatments

##### Chemotherapy

About one third (38.3%) of the study population received chemotherapy (Fig. S11A), and the rate of chemotherapy varied with age, BC subtype, and nodal status (Fig. S11B-D). Chemotherapy was administered in the neoadjuvant (*n*=15,627, 17.3%), adjuvant (*n*=72,939, 80.8%), or both settings (*n*=1,686, 1.9%) (Fig. S11E-H).

Chemotherapy regimens could be inferred in 55.0% of cases (*n*=50,559) but were unknown for 45.0% of cases (*n*=41,379) (Fig. 3A). The use of paclitaxel-only regimens increased with advancing age (Fig. 3B-C).

**Fig. 3:**
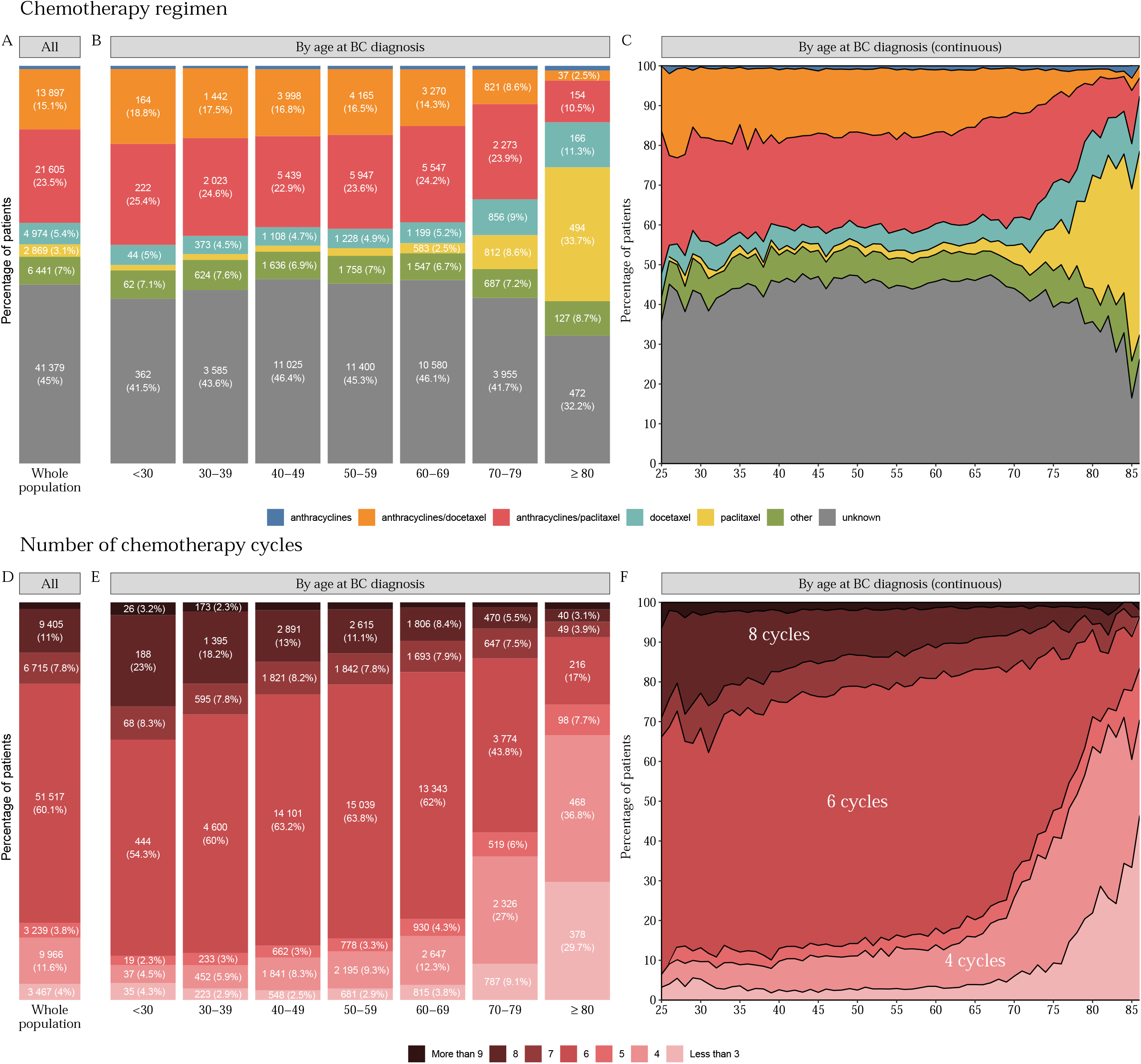
Chemotherapy regimen and number of cycles, by age at diagnosis. (A) Chemotherapy regimen for the total population (*n*=91,938). Raw data for subgroups representing less than 2% of the total population are not displayed on the graph, to ensure readability. In the anthracyclines group, *n*=773 (0.8%); (B) Chemotherapy regimen, by age class at BC diagnosis. Raw data for subgroups representing less than 2% of the corresponding age class are not displayed on the graph, to ensure readability. The values are: for the < 30 years old class: *n*=12 (1.4%), 30-39 years old: *n*=122 (1.5%), 40-49 years old: *n*=361 (1.5%), 50-59 years old: *n*=485 (1.9%) for the paclitaxel group. In the <30 years old class: *n*<10, 30-39 years old: *n*=49 (0.6%), 40-49 years old: *n*=203 (0.9%), 50-59 years old: *n*=205 (0.8%), 60-69 years old: *n*=202 (0.9%), 70-79 years old: *n*=90 (0.9%), ≥ 80 years old: 17 (1.2%) for the anthracyclines group; (C) Chemotherapy regimen, by age at BC diagnosis. The cohort is restricted to patients aged from 25 to 86 years (*n*=91,750); (D) Number of chemotherapy cycles for the total population. The number of chemotherapy cycles was calculated by setting (*n*=85,752). A patient with six cycles of neoadjuvant chemotherapy followed by four cycles of adjuvant chemotherapy was counted as having both six cycles and four cycles of treatment. Settings with missing numbers of chemotherapy cycles are not displayed (*n*=6,186). Raw data for subgroups representing less than 2% of the total population are not displayed on the graph, to ensure readability: 1,443 (1.7%) settings are in the more than 9 cycles group; (E) Number of chemotherapy cycles by age class (*n*=85,752) at BC diagnosis. Raw data for subgroups representing less than 2% of the corresponding age class are not displayed on the graph, to ensure readability. The values are: for the 40-49 years old class: *n*=441 (2.0%), 50-59 years old: *n*=412 (1.7%), 60-69 years old: *n*=271 (1.3%), 70-79 years old: *n*=98 (1.1%), ≥ 80 years old: *n*=22 (1.7%) for the more than 9 cycles group; (F) Chemotherapy regimens by age at BC diagnosis. The cohort is restricted to patients aged from 25 to 86 years (*n*=85,589). Abbreviations: BC = breast cancer, More than 9 = more than 9 cycles, 8 = 8 cycles, 7 = 7 cycles, 6 = 6 cycles, 5 = 5 cycles, 4 = 4 cycles, Less than 3 = less than 3 cycles.

Most patients received six cycles of chemotherapy (51,517, 60.1%), and the proportions of patients receiving four (9,966, 11.6%) or eight cycles (9,405, 11.0%) were similar (Fig. 3D). The number of cycles depended on age at BC diagnosis (Fig. 3E-F) and chemotherapy setting (Fig. S12A-F).

##### Targeted therapy

In total, 19,722 (8.4%) patients received targeted therapy, more frequently in the adjuvant setting than in the neoadjuvant followed by adjuvant setting (77.6% *versus* 22.4%, respectively; Fig. S13A-B) and more frequently with trastuzumab alone than with pertuzumab +/-trastuzumab (97.8% *versus* 2.2%, respectively; Fig. S13C-D).

TT was mostly combined with an anthracycline/docetaxel-based regimen (*n*=8,697, 44.1%) (Fig. S13E). The use of this regimen decreased with advancing age, in parallel with an increase in paclitaxel-TT (Tolaney) and TT associated with endocrine therapy (no chemotherapy) (Fig. S13F-G).

##### Endocrine therapy

In total, 165,655 patients (70.4%) received endocrine therapy, mostly in the adjuvant setting (Fig.4A-D). The three principal ET regimens were (1) AI (*n*=102,757, 62.0%), (2) tamoxifen (*n*=35,187, 21.2%) and (3) tamoxifen followed by AI (*n*=10,350, 6.2%), and the type of ET regimen depended strongly on age (Fig. 4F-G).

**Fig. 4:**
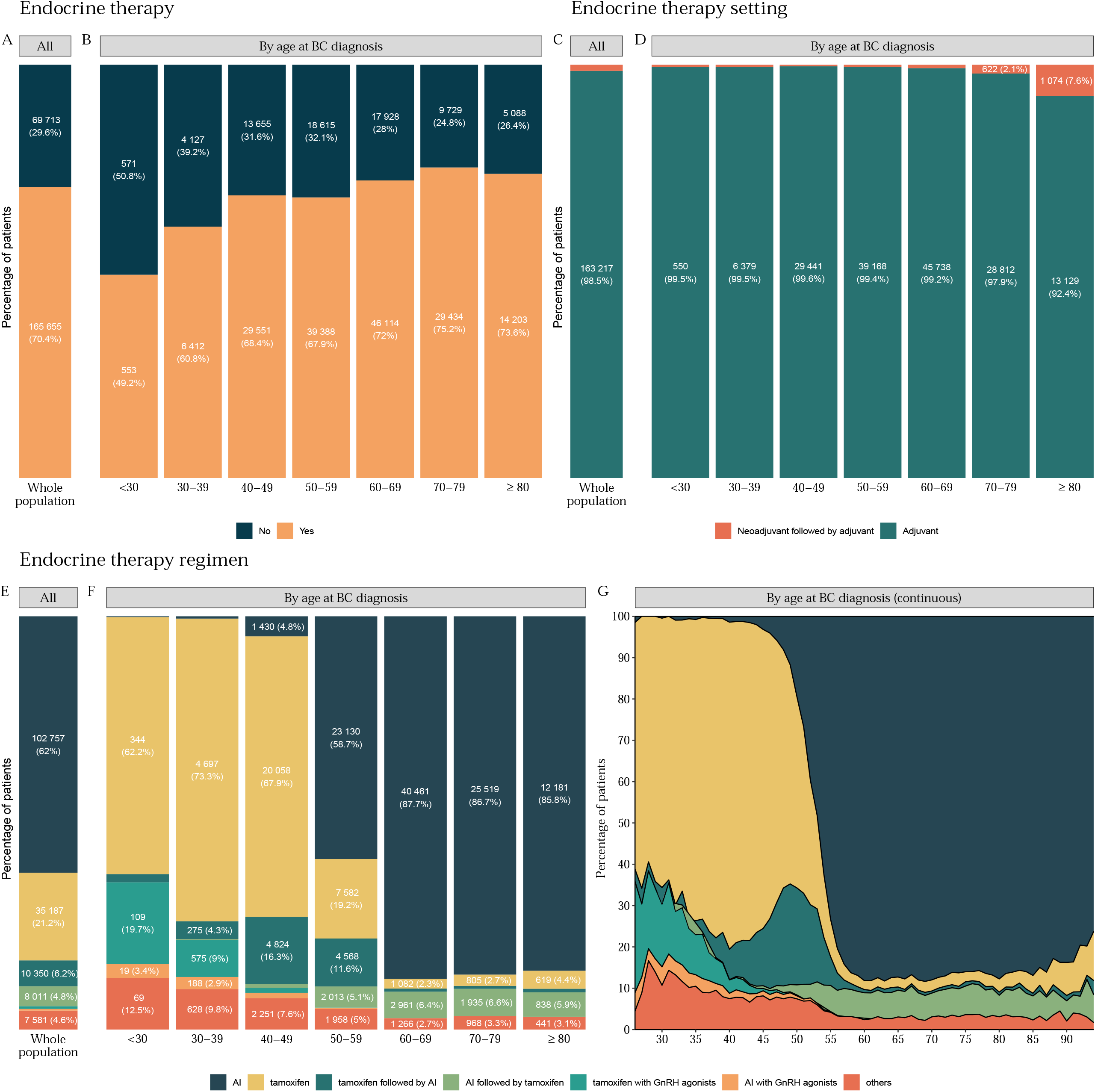
Endocrine therapy use, setting and regimens, by age at BC diagnosis. (A) Endocrine therapy in the total population (*n*=235,368); (B) Endocrine therapy by age class at BC diagnosis; (C) Endocrine therapy setting in total population (*n*=165,655). Raw data for subgroups representing less than 2% of the total population are not displayed on the graph, to ensure readability: there were 2,438 (1.5%) patients in the neoadjuvant followed by adjuvant group; (D) Endocrine therapy setting by age class at BC diagnosis. Raw data for subgroups representing less than 2% of the corresponding age class are not displayed on the graph, to ensure readability. The values not displayed are: for the <30 years old class: *n*<10, 30-39 years old: *n*=33 (0.5%), 40-49 years old: *n*=110 (0.4%), 50-59 years old: *n*=220 (0.6%), 60-69 years old: *n*=376 (0.8%) for the neoadjuvant followed by adjuvant group; (E) Endocrine therapy regimen for the total population (*n*=165,655). Raw data for subgroups representing less than 2% of the total population are not displayed on the graph, to ensure readability: there were 1,064 (0.6%) patients in the tamoxifen with GnRH agonists group, and 705 (0.4%) patients in the AI (=aromatase inhibitor) in combination with agonist group; (F) Endocrine therapy regimen by age class at BC diagnosis. Raw data for subgroups representing less than 2% of the corresponding age class are not displayed on the graph, to ensure readability.The values not displayed are: for the <30 years old group: *n*< 10, 30-39 years old: *n*=35 (0.5%) for the AI group. The values by age class for the tamoxifen followed by AI group are: for the < 30 years old group: *n*=11 (2%), 60-69 years old: *n*=341 (0.7%), 70-79 years old: *n*=207 (0.7%), ≥ 80 years old: *n*=124 (0.9%). The values by age class for the AI followed by tamoxifen group are: for the 30-39 years old group: *n*=14 (0.2%), 40-49 years old: *n*=250 (0.8%). The values by age class for the tamoxifen with GnRH agonists group are: for the 40-49 years old group: *n*=362 (1.2%), 50-59 years old: *n*=18 (0%). The values by age class for the AI with GnRH agonists group are: 40-49 years old: *n*= 376 (1.3%), 50-59 years old: *n*= 119 (0.3%), 60-69 years old: *n*<10. All other missing labels are 0; (G) Endocrine therapy regimen by age at BC diagnosis. The cohort is restricted to patients aged from 26 to 94 years (*n*=165,489); Abbreviations: BC= breast cancer, AI= aromatase inhibitor.

### Trends over time

The trends in treatments or pathways over time are displayed in Fig. S14. The proportion of patients undergoing a pathology procedure for diagnostic purposes before treatment increased over the study period (Fig. S14A-B). The type of breast surgery did not vary much over time (Fig. S14C-D). The proportion of patients treated with NAC increased slightly, the proportion of patients treated with adjuvant CT decreased, and the proportion of patients without CT increased (Fig. S14E-F). ET, CT, and TT combination regimens changed significantly over time (Fig. S14G-L).

### Survival outcomes

The median follow-up was 54.6 months (IQR: 33.9; 75.6), and 15,503 patients died (6.6%). Death was significantly associated with age at BC diagnosis (Fig. 5A), inferred BC subtype (Fig. 5B), and nodal status (Fig. 5C).

**Fig. 5:**
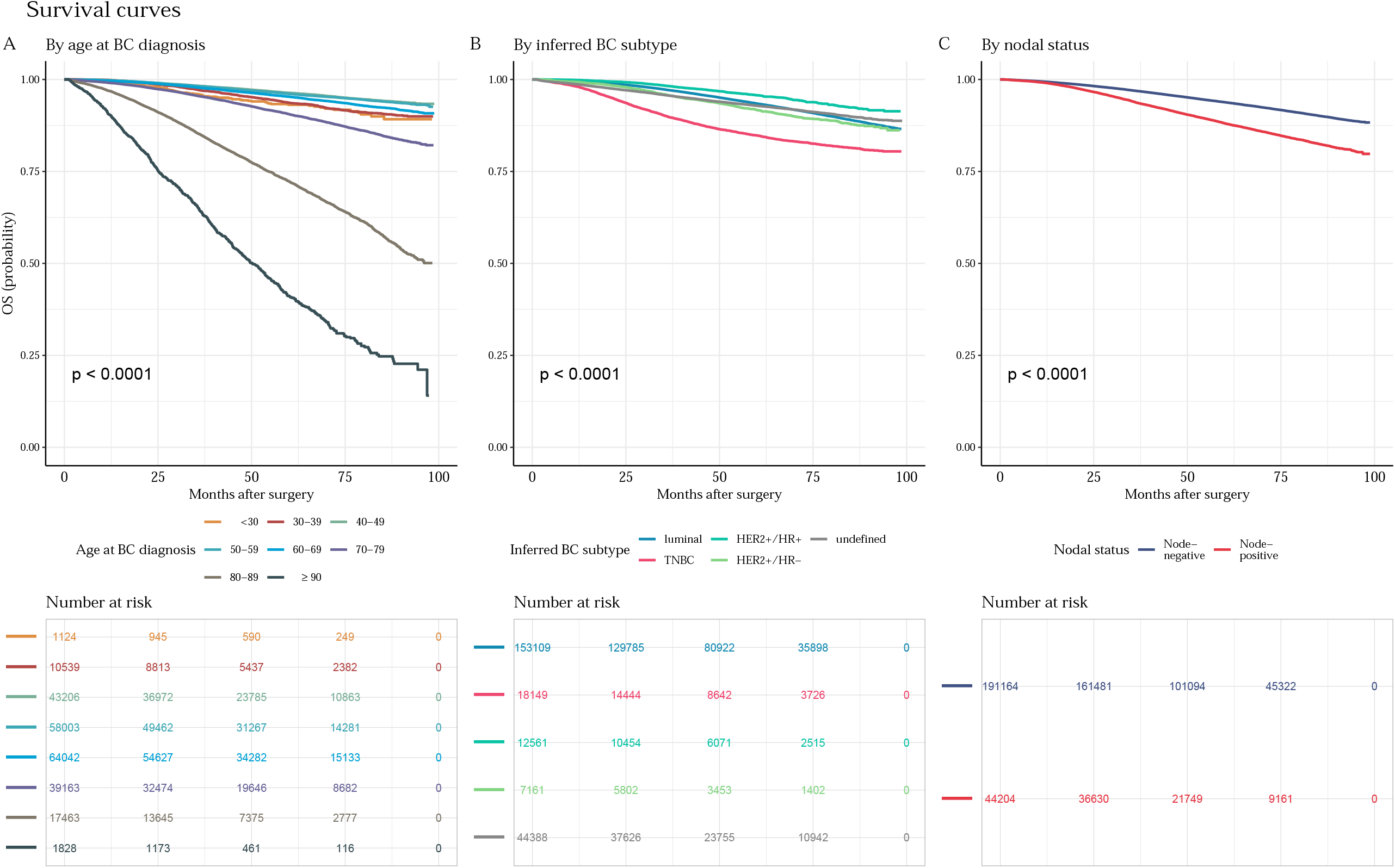
Unadjusted Kaplan-Meier survival curves. (A) Unadjusted Kaplan-Meier survival curves for the association between overall survival (OS) and age class at BC diagnosis. (B) Unadjusted Kaplan-Meier survival curves for the association between OS and inferred BC subtype. (C) Unadjusted Kaplan-Meier survival curves for the association between OS and nodal status.

## Discussion

In this study of 235,368 French women newly diagnosed with early BC, we constitute one of the largest national cohorts of BC patients treated within a universal healthcare system described to date. This resource will be very useful for a number of reasons.

First, the SNDS is one of the largest exhaustive nationwide aggregated health data resources worldwide (Moulis et al., 2015b). Several other large databases of BC patients exist worldwide. The Surveillance, Epidemiology, and End Results (SEER) program of the National Cancer Institute collects data on cancer diagnoses, treatment, and survival for approximately 30% of the United States (US) population (Duggan et al., 2016). The National Cancer Database (NCDB) has amassed more than 34 million hospital records for cancer patients (Boffa et al., 2017) and contains data for patients from the United States who received any element of their cancer care as part of a cancer program accredited by the American College of Surgeons Commission of Cancer (CoC). The NCCN Breast Cancer Outcomes Database (DB) contains data for patients receiving all or some of their treatment at one of eight US reporting centers (Punglia et al., 2008; Vaz-Luis et al., 2014). The limitations of these databases include a lack of exhaustivity. There may also be biases, as NCDB and NCCN Breast Cancer Outcomes DB are not population-based databases and only consider the care of those who had access to and received treatment at major academic cancer centers. Such biases can be ruled out in the exhaustive population-based registries of northern European countries such as Sweden, Norway, Finland and Denmark (de Boniface et al., 2021; Christiansen et al., 2016; Johansson et al., 2021; Leinonen et al., 2017). We report here a much larger cohort of BC patients, which could be used for studies with high statistical power.

One major challenge in the use of reimbursement databases is the accuracy with which the reimbursement indicators reflect the medical condition. Over the last decade, the accuracy and reliability of SNDS have been evaluated for a very large range of medical conditions, in patients with prosthetic heart valves (Tubiana et al., 2017), in patients with Crohn’s disease (Meyer et al., 2019), in parturient women (Boucheron et al., 2021), or for risks associated with exposure to certain treatments, such as cyproterone acetate (Weill et al., 2021), statins (Giral et al., 2019), thiopurines or TNF antagonists (Lemaitre et al., 2017). Several studies have been performed in the BC field. Algorithms for identifying incident cancer cases in French administrative health databases have been published (Ajrouche et al., 2017) (Bousquet et al., 2017). Other studies have described the care pathways of BC patients (Lefeuvre et al., 2017, 2019), compliance with endocrine therapy (Huiart et al., 2012; Lailler et al., 2021) or the risk of hematologic malignancies following the use of G-CSF (Jabagi et al., 2020). There have also been studies focusing on particular conditions in specific subpopulations, such as BC in male patients (Cottenet et al., 2019).

In our cohort, the distributions of the proxies for BC we describe are consistent with those from previous studies. We excluded 4.5% of the population due to a suspicion of stage IV BC at diagnosis, consistent with published rates, which are usually estimated at about 5% (DeSantis et al., 2019)(Daily et al., 2021). The rate of nodal involvement was also similar to the value of about 25% for the SEER data (Dreyer et al., 2018), and the increase in risk from luminal to TNBC to *HER2*^*+*^ BCs was also similar (Yang et al., 2017). We also report decreasing proportions of TNBC and *HER2*^+^ BCs with advancing age, consistent with epidemiological evidence (Murphy et al., 2019)(Kim et al., 2022). Our data are also consistent with the trends in clinical practice over time. The proportion of paclitaxel – trastuzumab regimens increased significantly after 2015, following the publication of Tolaney’s work (Tolaney et al., 2015), which introduces the use of adjuvant paclitaxel plus trastuzumab in small node-negative *HER2*^+^ BCs. Similarly, seven fatal cases of toxic enterocolitis occurred in France in 2016 and were suspected to be linked to docetaxel. In response, docetaxel was temporarily banned following warnings issues by the French national agency for medicines and health product safety (ANSM), and our data show a dramatic increase in the use of paclitaxel-based CT regimens as an alternative to docetaxel since January 2017.

The strengths of our study include the collection of data for an unprecedented number of BC patients. The use of reimbursement data is particularly appropriate in this setting because inferences can be made about BC biology due to the specificity of treatments, such as endocrine therapy and anti-*HER2* therapies targeting particular molecular alterations.

Conversely, we cannot exclude the possibility of a selection bias because we applied stringent criteria for patient inclusion, to ensure that only patients with high-quality data were retained. The identification of patterns of treatment based on codes (ICD-10, CCAM etc.) may also be subject to coding errors or discrepancies in coding methods between centers. Finally, BC subtypes were defined on the basis of treatment, making it impossible to classify patients with HR^+^ cancers who refused ET or patients with *HER2*^+^ BCs that were not treated with targeted therapies.

The FRESH cohort opens up multiple perspectives. At patient level, such a resource could be used to analyze outpatient care during and after treatment, to identify rare adverse events, or to monitor early complications of treatment or patterns of late sequalae. At cancer care center level, benchmarks could be established with performance or quality indicators, such as EUSOMA (Biganzoli et al., 2017), and real-time monitoring of care could be implemented in the context of continually changing guidelines. At national level and from an economic standpoint, this cohort could be used to perform medico-economic studies, and to rationalize healthcare expenditure. Finally, in terms of research and development, this cohort represents a strategic opportunity for generating hypotheses and optimizing the potential for innovation to improve cancer care.

## Data Availability

All data produced in the present study are available upon reasonable request to the authors.

## Acknowledgments

We thank the Department of Health Data and Assessment, Survey Data Science and Assessment Division, French National Cancer Institute (Institut National du Cancer INCa) for providing us with access to the cancer cohort.

---

**Table S1: ICD-10 diagnosis codes used to identify breast cancer**

Abbreviations: ICD 10 = International Statistical Classification and Related Health Problems 10th revision.

**Table S2: ICD-10 and ATC codes used to identify cancer at another site**.

Abbreviations: ICD-10 = International Statistical Classification and Related Health Problems 10th revision; ATC = Anatomical Therapeutic and Chemical Classification.

**Table S3: ICD-10 and ATC codes used to identify *de novo* metastatic breast cancer**

Abbreviations: ICD 10 = International Statistical Classification and Related Health Problems 10th revision; ATC = Anatomical Therapeutic and Chemical Classification; *HER2*=human epidermal growth factor receptor 2.

**Table S4: CCAM, ICD-10 and ATC codes used to identify breast cancer treatments**. Abbreviations: CCAM = French Common Classification of Medical Procedures; ICD 10 = International Statistical Classification and Related Health Problems – 10th revision; ATC = Anatomical Therapeutic and Chemical Classification; *HER2*=human epidermal growth factor receptor 2.

**Table S5: CCAM, ATC, CIP and UCD codes used to identify embryo or oocyte cryopreservation**

Abbreviations: CCAM = French Common Classification of Medical Procedures; ATC = Anatomical Therapeutic and Chemical Classification; CIP (*Code Identifiant de Présentation*); UCD (*Unités Communes de Dispensation*).

**Table S6: CCAM codes used to identify diagnostic procedures for breast cancer** Abbreviations: CCAM = French Common Classification of Medical Procedures; CT = computed tomography.

**Fig. S1: Flowchart of the French Early Breast Cancer Cohort**.

Abbreviations: BC = breast cancer.

**Fig. S2**: **Rules for the identification of chemotherapy regimens and the number of cycles of chemotherapy in patients not treated with targeted therapy**.

The detailed methodology for identifying chemotherapy regimens is described in the Supplementary Material. *n* is the number of seven-day intervals between chemotherapy sessions; *q* is the number of 14-day intervals between chemotherapy sessions; *p* is the number of 21-day intervals between chemotherapy sessions; and E() is the floor function.

**Fig. S3**: **Rules for the identification of chemotherapy regimens, combinations of targeted therapy and systemic treatment regimens, numbers of cycles of chemotherapy and numbers of cycles of targeted therapy in patients treated with both chemotherapy and targeted therapy**.

The detailed methodology for identifying chemotherapy regimens is described in the Supplementary Material. *n* is the number of seven-day intervals between chemotherapy sessions; *q* is the number of 14-day intervals between chemotherapy sessions; *p* is the number of 21-day intervals between chemotherapy sessions; and E() is the floor function.

**Fig. S4: Age at BC diagnosis, by year of BC diagnosis**

Distribution of age at diagnosis by year of diagnosis. The hypothesis of unimodality was significantly rejected for all years of diagnosis (*p*<0.001) and we determined visually that each distribution had two modes. The inferred modes are displayed on the graph.

**Fig. S5: Mode of diagnosis, by age at BC diagnosis**

(A) Modes of diagnosis for the total population (*n*=235,368). Raw data for subgroups representing less than 2% of the whole population are not displayed on the graph, to ensure readability: there were 2,436 (1%) patients in the no diagnostic procedure group;

(B) Mode of diagnosis by age class at BC diagnosis. Raw data for subgroups representing less than 2% of the corresponding age class are not displayed on the graph, to ensure readability. The values per age class not displayed are: 30-39 years old: *n*=168 (1.6%), 40-49 years old: *n*=401 (0.9%), 50-59 years old: *n*=408 (0.7%), 60-69 years old: *n*=397 (0.6%), 70-79 years old: *n*=337 (0.9%) for the no diagnostic procedure group. The values by age group for the cytology group are: 40-49 years old: *n*=857 (2%), 50-59 years old: 1,121 (1.9%), 60-69 years old: *n*= 1,239 (1.9%);

(C) Mode of diagnosis by age at BC diagnosis. The cohort is restricted to patients aged from 24 to 95 years (*n*=235,152);

(D) Modes of diagnosis in patients with surgery as the first treatment, for the total population (*n*=215,701). Raw data for subgroups representing less than 2% of the whole population are not displayed on the graph, to ensure readability: 2,007 (0.9%) patients had no diagnostic procedure;

(E) Modes of diagnosis for patients with surgery as the first treatment, by age class at BC diagnosis. Raw data for subgroups representing less than 2% of the corresponding age class are not displayed on the graph, to ensure readability. The values by age class not shown are: 30-39 years old: *n*=109 (1.4%), 40-49 years old: *n*=308 (0.8%), 50-59 years old: *n*=339 (0.6%), 60-69 years old: *n*=305 (0.5%), 70-79 years old: *n*=287 (0.8%) for the no diagnostic procedure group. The values by age class for the cytology group are: 50-59 years old: 1,054 (2.0%), 60-69 years old: *n*=1,177 (1.9%);

(F) Modes of diagnosis for patients with surgery as the first treatment, by age at BC diagnosis. The cohort is restricted to patients aged from 25 to 95 years (*n*=215,464);

(G) Modes of diagnosis in patients with neoadjuvant treatment in the total population (*n*=17,479). Raw data for subgroups representing less than 2% of the total population are not displayed on the graph, to ensure readability: there were 323 (1.8%) patients in the no diagnostic procedure group, and 306 (1.8%) in the cytology group;

(H) Modes of diagnosis in patients with neoadjuvant treatment by age class at BC diagnosis. Raw data for subgroups representing less than 2% of the corresponding age class are not displayed on the graph, to ensure readability. The values by age class not shown are: <30 years old: <10, 40-49 years old: 85 (1.6%), 50-59 years old: 61 (1.3%) for the no diagnostic procedure group. The values by age class for the cytology group are: < 30 years old: *n* < 10, 40-49 years old: *n*=90 (1.7%), 50-59 years old: *n*=67 (1.5%), 60-69 years old: *n*=59 (1.9%), 70-79 years old: *n*=21 (1.8%);

(I) Modes of diagnosis in patients with neoadjuvant treatment, by age at BC diagnosis. The cohort is restricted to patients aged from 27 to 79 years (*n*=17,132).

Abbreviations: BC = breast cancer.

**Fig. S6: Inferred BC subtype, including undefined tumors, by age at BC diagnosis**

(A) Inferred BC subtype for the total population (*n*=235,268);

(B) Inferred BC subtype by age class at BC diagnosis; raw data for subgroups representing less than 2% of the corresponding age class are not displayed on the graph, to ensure readability: ≥ 80 years old: 325 (1.7%) patients in the *HER2*^*+*^/HR^+^ group and 259 (1.3%) patients in the *HER2*^*+*^/HR^−^ group.

(C) Inferred BC subtype by age at BC diagnosis. The cohort is restricted to patients aged from 24 to 95 years with an undefined subtype (*n*=235,152).

Abbreviations: BC = breast cancer; HR^+^ = hormone receptor-positive; HR^−^ = hormone receptor-negative; TNBC = triple-negative breast cancer subtype; *HER2*^*+*^ = human epidermal growth factor receptor 2.

**Fig. S7: Age at BC diagnosis, for undefined tumors**

Age distribution of patients with undefined, inferred subtypes (*n*=44,388) at BC diagnosis (*p*-value for non-unimodality < 0.001).

Abbreviations: BC = breast cancer.

**Fig. S8**: **Nodal status by age at BC diagnosis and inferred BC subtype**

(A) Nodal status in the total population (*n*=235,368);

(B) Nodal status per age class at BC diagnosis;

(C) Nodal status per inferred BC subtype.

**Fig. S9: Type of surgery, by age at BC diagnosis, inferred BC subtype and nodal status**.

(A) Breast surgery type in the total population (*n*=235,368);

(B) Breast surgery type by age class at BC diagnosis;

(C) Breast surgery type by inferred BC subtype;

(D) Breast surgery type by nodal status at diagnosis;

(E) Use of axillary surgery in the total population (*n*=235,368);

(F) Use of axillary surgery by age class at BC diagnosis;

(G) Use of axillary surgery by inferred BC subtype;

(H) Use of axillary surgery by nodal status at diagnosis.

Abbreviations: BC = breast cancer; TNBC = triple-negative breast cancer subtype; *HER2*^*+*^ = human epidermal growth factor receptor 2; HR^+^ = hormone receptor-positive; HR^−^ = hormone receptor-negative.

**Fig. S10: Radiotherapy by age at BC diagnosis, inferred BC subtype and nodal status**.

(A) Radiotherapy in the total population (*n*=235,368);

(B) Radiotherapy by age class at BC diagnosis;

(C) Radiotherapy by inferred BC subtype;

(D) Radiotherapy by nodal status at diagnosis;

(E) Radiotherapy in patients treated by partial mastectomy (*n*=173,173);

(F) Radiotherapy in patients treated by partial mastectomy, by age class at BC diagnosis;

(G) Radiotherapy in patients treated by partial mastectomy, by inferred BC subtype. Raw data for subgroups representing less than 2% of the corresponding class are not displayed on the graph, to ensure readability: in the *HER2*^*+*^/HR^+^ category, 132 (1.6%) patients did not receive radiotherapy.

(H) Radiotherapy in patients treated by partial mastectomy, by nodal status at diagnosis.

Abbreviations: BC = breast cancer; HR^+^ = hormone receptor-positive; HR^−^ = hormone receptor-negative; *HER2*^*+*^ = human epidermal growth factor receptor 2; TNBC = triple-negative breast cancer subtype.

**Fig. S11: Chemotherapy use and setting by age at BC diagnosis, inferred BC subtype and nodal status**

(A) Use of chemotherapy in the total population (*n*=235,368);

(B) Use of chemotherapy per age class at BC diagnosis;

(C) Use of chemotherapy per inferred BC subtype; Raw data for subgroups representing less than 2% of the corresponding breast subtype class are not displayed on the graph, to ensure readability: in the *HER2*^*+*^/HR^+^ category, there were 137 (1.1%) patients in the no chemotherapy group;

(D) Use of chemotherapy by nodal status;

(E) Chemotherapy setting in the total population (*n*=90,252). Raw data for subgroups representing less than 2% of the total population are not displayed on the graph, to ensure readability: there were 1,686 (1.9%) patients in the “both” category;

(F) Chemotherapy setting by age class at BC diagnosis. Raw data for subgroups representing less than 2% of the corresponding age class are not displayed on the graph, to ensure readability. The values by age class are: 50-59 years old: *n*=449 (1.8%), 60-69 years old: *n*=292 (1.3%), 70-79 years old: *n*=96 (1.0%), ≥ 80 years old: *n*=23 (1.6%) for the “both” group;

(G) Chemotherapy setting by inferred BC subtype; raw data for subgroups representing less than 2% of the corresponding breast subtype class are not displayed on the graph, to ensure readability: in the luminal category, there were 454 (0.9%) patients in the “both” group;

(H) Chemotherapy setting by nodal status; Raw data for subgroups representing less than 2% of the corresponding nodal status group are not displayed on the graph, to ensure readability: in the node-positive category, there were 500 (1.6%) patients in the “both” group; Abbreviations: BC = breast cancer, HR^+^ = hormone receptor-positive; HR^−^ = hormone receptor-negative; *HER2*^*+*^=human epidermal growth factor receptor 2; TNBC = triple-negative breast cancer subtype.

**Fig. S12: Number of chemotherapy cycles by age at BC diagnosis in the neoadjuvant and adjuvant settings**

(A) Number of neoadjuvant chemotherapy cycles for the total population. The number of neoadjuvant chemotherapy cycles was calculated by setting (*n*=16,106). Settings with missing numbers of chemotherapy cycles are not displayed (*n*=1,207). A patient with six cycles of neoadjuvant chemotherapy followed by four cycles of adjuvant chemotherapy was classified as having six cycles in subfigures A-B-C; and four in subfigures D-E.-F;

(B) Number of neoadjuvant chemotherapy cycles by age class (*n*=16,106) at BC diagnosis. Raw data for subgroups representing less than 2% of the corresponding age class are not displayed on the graph, to ensure readability. The values by age class are: 60-69 years old: *n*=47 (1.6%), 70-79 years old: *n*=20 (2.0%), ≥ 80 years old: *n*<10 for the more than 9 cycles group;

(C) Number of neoadjuvant chemotherapy cycles by age at BC diagnosis. The cohort is restricted to patients aged from 27 to 79 years (*n*=15,810);

(D) Number of adjuvant chemotherapy cycles for the total population (*n*= 69,646). Settings with missing numbers of chemotherapy cycles are not displayed (*n*=4,979). Raw data for subgroups representing less than 2% of the total population are not displayed on the graph, to ensure readability: there were 1,044 (1.5%) patients in the more than 9 cycles group;

(E) Number of adjuvant chemotherapy cycles by age class (*n*=69,646) at BC diagnosis. Raw data for subgroups representing less than 2% of the corresponding age class are not displayed on the graph, to ensure readability. The values by age class are: 30-39 years old: *n*=93 (1.8%), 40-49 years old: *n*=303 (1.7%), 50-59 years old: *n*=315 (1.6%), 60-69 years old: *n*=224 (1.2%), 70-79 years old: *n*=78 (1.0%), ≥ 80 years old: *n*=19 (1.8%) for the more than 9 cycles group;

(F) Number of adjuvant chemotherapy cycles by age at BC diagnosis. The cohort is restricted to patients aged from 26 to 85 years (*n*=69,452).

Abbreviations: BC = breast cancer, More than 9 = more than 9 cycles, 8 = 8 cycles, 7 = 7 cycles, 6 = 6 cycles, 5 = 5 cycles, 4 = 4 cycles, Less than 3 = less than 3 cycles.

**Fig. S13: Targeted therapy setting and regimen by age at BC diagnosis**

(A) Targeted therapy settings for the total population (*n*=19,722);

(B) Targeted therapy setting by age class at BC diagnosis;

(C) Targeted therapy regimens for the total population (*n*=19,722);

(D) Targeted therapy regimen by age class at BC diagnosis. There were 433 (2.2%) patients in the pertuzumab +/-trastuzumab category, three of whom received pertuzumab only, the remaining 430 being treated with both pertuzumab and trastuzumab. Raw data for subgroups representing less than 2% of the corresponding age class are not displayed on the graph, to ensure readability. The values by age class are: 50-59 years old: *n*=102 (1.8%), 60-69 years old: *n*=92 (2.0%), 70-79 years old: *n*= 39 (1.8%) for the pertuzumab +/-trastuzumab group;

(E) Combinations of TT and systemic treatment regimens for the total population (*n*=19,722). There were 190 (1.0%) patients in the targeted therapy-endocrine therapy group;

(F) Combination of TT and systemic treatment regimen by age class at BC diagnosis. Raw data for subgroups representing less than 2% of the corresponding age class are not displayed on the graph, to ensure readability. The values by age class are: < 30 years old: *n*<10, 30-39 years old: *n*<10, 40-49 years old: *n*=20 (0.4%), 50-59 years old: *n*=32 (0.6%), 60-69 years old: *n*=39 (0.8%), 70-79 years old: *n*=35 (1.6%) for the TT-ET (no chemotherapy) group. In the < 30 years old category, < 10 patients were in the paclitaxel-TT (Tolaney) group.

(G) Combination of TT and systemic treatment regimen by age at BC diagnosis. The cohort is restricted to patients aged from 28 to 84 years (*n*=19,487).

Abbreviations: BC = breast cancer, TT = targeted therapy, ET = endocrine therapy.

**Fig. S14: Changes in medical practices over time**

(A) Mode of diagnosis by year of diagnosis. Raw data for subgroups representing less than 2% of the corresponding year class are not displayed on the graph, to ensure readability. The values by year class are: in 2014: *n*=659 (1.9%), 2015: *n*=514 (1.5%), 2016: *n*=553 (1.6%), 2017: *n*=490 (1.4%) for the cytology group. The values by year class for the no diagnostic procedure group are: in 2011: *n*=409 (1.3%), 2012: *n*=356 (1.1%), 2013: *n*=362 (1.1%), 2014: *n*=345 (1.0%), 2015: *n*=348 (1.0%), 2016: *n*=308 (0.9%), 2017: *n*=308 (0.9%);

(B) Mode of diagnosis by month-year of diagnosis (*n* = 235,368);

(C) Type of breast surgery by year of diagnosis;

(D) Type of breast surgery by month-year of diagnosis (*n* = 235,368);

(E) Main care pathways by year of diagnosis. Raw data for subgroups representing less than 2% of the corresponding year class are not displayed on the graph, to ensure readability. The values by year class are: in 2011: *n*=331 (1.0%), 2012: *n*=306 (0.9%), 2013: *n*=291 (0.9%), 2014: *n*=289 (0.9%), 2015: *n*=298 (0.9%), 2016: *n*=352 (1.0%), 2017: *n*=321 (0.9%) for the NET group;

(F) Main care pathways by month-year of diagnosis (*n*= 235,202). Patients who received neoadjuvant radiotherapy but not neoadjuvant chemotherapy are classified as NRT. They are not displayed on the plot (*n* =166).

(G) Endocrine therapy regimen by year of diagnosis. Raw data for subgroups representing less than 2% of the corresponding year class are not displayed on the graph, to ensure readability. The values by year class are: in 2011: *n*=31 (0.1%), 2012: *n*=42 (0.2%), 2013: *n*=52 (0.2%), 2014: *n*= 86 (0.4%), 2015: *n*=133 (0.6%), 2016: *n*=167 (0.7%), 2017: *n*=194 (0.8%) for the AI with GnRH agonists group. The values by year class are: in 2011: *n*=143 (0.6%), 2012: *n*=148 (0.6%), 2013: *n*=124 (0.5%), 2014: *n*=157 (0.5%), 2015: *n*=155 (0.6%), 2016: *n*=165 (0.7%), 2017: *n*=172 (0.7%) for the tamoxifen with GnRH agonists group. In the 2017 category, there were 338 (1.4%) patients in the tamoxifen followed by AI group and 271 (1.1%) patients in “others” group;

(H) Endocrine therapy regimen by month-year of ET start date. The cohort is restricted to patients whose month-year ET start date was after March 2011 and before November 2018 (*n* = 165,569);

(I) Chemotherapy regimen by year of diagnosis. Raw data for subgroups representing less than 2% of the corresponding year class are not displayed on the graph, to ensure readability: in the 2012 category, there were 16 (0.1%) patients in the anthracyclines group and 229 (1.7%) in the paclitaxel group; in the 2011 and 2013 categories, there were 194 (1.5%) and 244 (1.8%) patients, respectively, in the paclitaxel group;

(J) Chemotherapy regimen by month-year of CT start date. The cohort is restricted to patients whose month-year of CT start date was after February 2011 and before May 2018 (*n* = 95,175);

(K) Combinations of TT and other systemic treatments by year of diagnosis. Raw data for subgroups representing less than 2% of the corresponding year class are not displayed on the graph, to ensure readability. The values by year class are: in 2011: *n*=24 (1.0%), 2012: *n*=24 (1.0%), 2013: *n*=34 (1.3%), 2014: *n*=41 (1.4%), 2015: *n*=30 (1.0%), 2016: *n*=16 (0.5%), 2017: *n*=21 (0.7%) for the targeted therapy - endocrine therapy (no chemotherapy) group;

(L) Combinations of TT and other systemic treatments by month-year of TT start date. The cohort is restricted to patients whose month-year of TT start date was after April 2011 and before May 2018 (*n* = 19,658).

Abbreviations: BC = breast cancer, CT = chemotherapy, NAC = neoadjuvant chemotherapy; NET = neoadjuvant endocrine therapy, TT = targeted therapy, ET = endocrine therapy, AI = aromatase inhibitor.

