## Supplemental material for "The French Early Breast Cancer Cohort (FRESH): a resource for breast cancer research and evaluations of oncology practices based on the French National Healthcare System Database (SNDS)"

***Patient selection***

We applied 10 filters for patient selection (Fig. S1):

- (1) BC diagnosis was identified by a tag with an ICD-10 diagnosis code for BC (Table S1) in long-term illness (LTI) records; or in at least one hospital discharge report within the period considered. The inclusion year was defined as the year of the first diagnosis code for BC.
- (2) Patients were considered to have BC newly diagnosed between January 1, 2011 and December 31, 2017 if the inclusion year was between 2011 and 2017. Patients with a BC diagnosis code in 2010 were excluded from the analyses.
- (3) Female patients were identified from the binary sex variable directly available in ODP. Male patients were excluded from the cohort.
- (4) Age at BC diagnosis was calculated as the rounded difference, in years, between the date of BC diagnosis and the patient's date of birth. Patients under 18 years of age at BC diagnosis or whose date of birth was missing in the ODP were excluded.
- (5) Registration to general health insurance coverage plan ("Régime Général") was identified from the insurance scheme table available in ODP, which summarizes the start and end date of the patient's insurance scheme. Patients affiliated to another insurance scheme or with a change of affiliation between the inclusion year and 2018 were excluded.
- (6) BC surgery was identified as described in the surgery section. Women with no breast surgery at any time in the inclusion year or the following year were excluded from the

study. We defined BC index surgery as the first breast surgery intervention occurring in the inclusion year or the following.

- (7) Patients suspected to have been diagnosed with BC at the same time as cancer at another site were excluded. These patients were identified by at least one diagnosis code (hospital discharge or LTI) for the other cancer (Table S2), except for cervical intraepithelial neoplasia and basal or squamous cell carcinoma of skin, or by the administration of chemotherapy or immunotherapy molecules not indicated for BC (Table S2) in the year preceding or following BC index surgery.
- (8) We excluded diagnoses of BC presenting as locoregional or distant relapses from a previous cancer diagnosed before the study period, by excluding patients with an LTI for BC starting before January 1, 2011, or a diagnosis code for a prior BC (ICD-10 Z853) before or up to six months after BC index surgery.
- (9) Stage IV BC at diagnosis was suspected on the basis of at least one diagnosis code for metastatic disease (Table S3); or at least one hospital administration or outpatient delivery of chemotherapy, targeted therapy or endocrine therapy molecules indicated only for metastatic disease (Table S3); or at least three days of daily chemotherapy sessions before or up to six months (as previously described, (Grinda et al., 2021)) after BC index surgery; or more than 20 cycles of anti-*HER2* targeted therapy. The corresponding patients were excluded from the study population. The methodology used to identify daily chemotherapy sessions and the number of cycles of anti-*HER2* targeted therapy is detailed below.
- (10) Data quality was checked to exclude patients with missing data for area of residence, inconsistent numbers of chemotherapy sessions within a hospital stay or a refined date of diagnosis in 2010 or 2018, based on treatment dates and biopsy procedure dates.

***BC treatments***

**Surgery**

Starting with the BC index surgery and then following successive hospitalizations, the surgical procedures occurring after the BC index surgery were considered to be revision surgery interventions provided that (i) they occurred within three months of the previous surgical intervention, (ii) they constituted the first surgical procedure after adjuvant chemotherapy, less than three months after the last chemotherapy session. The methodology used to identify adjuvant chemotherapy is detailed below. Final binning for breast surgery into two categories was as follows: (1) *mastectomy,* if at least one operation from among the BC index surgery and revision surgery interventions was mastectomy, with or without axillary surgery; (2) *partial mastectomy* otherwise. Final binning for axillary surgery into two categories was as follows: (1) *Yes*; if at least one of the surgical interventions from among the BC index surgery and the revision surgery operations was mastectomy with axillary surgery, partial mastectomy with axillary surgery or axillary surgery without breast surgery; (2) *No*; otherwise.

**Radiotherapy**

Radiotherapy (RT) sessions were tagged as neoadjuvant if they occurred before BC index surgery and adjuvant otherwise.

Radiotherapy settings were tagged as: (1) *neoadjuvant* if all RT sessions were tagged as neoadjuvant; (2) *adjuvant* if all RT sessions were tagged as adjuvant; (3) *both* if at least one RT session was tagged as neoadjuvant and at least one RT session was tagged as adjuvant.

The neoadjuvant *RT start date* was defined as the date of the first session of neoadjuvant RT between 150 days before BC index surgery and BC index surgery. The neoadjuvant *RT end date* was defined as the date of the last session of neoadjuvant radiotherapy within 150 days of the neoadjuvant RT start date.

The adjuvant *RT start date* was defined as the date of the first session of adjuvant RT from BC index surgery to 365 days after BC index surgery. The adjuvant *RT end date* was defined as the date of the last session of adjuvant radiotherapy within 150 days of the adjuvant RT start date.

**Chemotherapy**

*Chemotherapy setting and calculation of chemotherapy dates*

CT sessions were tagged as neoadjuvant if they occurred before the BC index surgery date and adjuvant otherwise.

CT settings were tagged as: (1) *neoadjuvant* if all CT sessions were tagged as neoadjuvant; (2) *adjuvant* if all CT sessions were tagged as adjuvant; (3) *both* if at least one CT session was tagged as neoadjuvant and at least one CT session was tagged as adjuvant.

The neoadjuvant *CT start date* was defined as the date of the first session of neoadjuvant CT between 250 days before BC index surgery and BC index surgery. The neoadjuvant *CT end date* was defined as the date of the last session of neoadjuvant CT within either seven months of the neoadjuvant CT start date or the first date of suspicion of metastatic progression on treatment (administration of molecules indicated only for metastatic disease listed in Table S3 or at least three consecutive daily chemotherapy sessions), whichever occurred first.

The adjuvant *CT start date* was defined as the date of the first session of adjuvant CT between BC index surgery to 180 days after BC index surgery. The adjuvant *CT end date* was defined as the date of the last session of adjuvant CT within either seven months of the adjuvant CT start date or the first date of suspicion of metastatic progression on treatment (administration of molecules indicated only for metastatic disease listed in Table S3 or at least three consecutive daily chemotherapy sessions), whichever occurred first.

*Computation of intervals between chemotherapy sessions*

Hospital stays (outpatient care or long-term hospitalizations) including CT sessions were characterized by their start date, end date and the number of CT sessions performed. Patients with any hospital stay with an inconsistent number of CT sessions (*i.e.* more than 1 session per day) were excluded from the cohort. We then distinguished two categories of patients.

- The first category included patients for whom CT sessions were all performed in the day hospital (hospital outpatient care), such that the exact date of each CT session was known. Most of the patients (96.7%) belonged to this category. For these patients, the intervals between two consecutives CT sessions were calculated as the difference in CT session dates, rounded to 1, 7, 14 or 21 days.
- The second category included patients for whom at least one CT session date was unknown, making it impossible to calculate directly all the intervals between consecutive CT sessions. Only a minority of patients belonged to this category (3.3 %). We inferred the sequence of CT session intervals for these patients from the most frequent sequence of intervals for patients in the first category that matched the available data for the timing of CT sessions for the patient concerned. If the inferred sequence of

intervals was not among the 20 most frequent sequences for patients in the first category, the inference was considered uncertain and the patient was excluded from the cohort.

Daily chemotherapy sessions (sessions separated by an interval of one day) were tagged as indicating a suspicion of metastatic disease. If at least three consecutive daily CT sessions were identified within six months of the BC index surgery, the patient was considered to have had stage IV BC at diagnosis and was excluded from the cohort. If at least three consecutive daily CT sessions were identified six months or more after BC index surgery, the patient was assumed to have distant metastases or disease progression, and all subsequent CT sessions were excluded from calculation of the CT regimen.

*Identification of CT regimen and the number of cycles*

Apart from costly innovative drugs part of a special reimbursement process in France called "list en sus"), chemotherapy molecules were not directly identifiable in hospital care. Nevertheless, chemotherapy regimen may still sometimes be inferred from specific temporal schemes (*e.g.* paclitaxel is the only early BC chemotherapy molecules to be delivered weekly). The decision rules applied to patients without targeted therapies are displayed in Fig. S2:

(1) Any succession of seven-day intervals was considered to identify a paclitaxel regimen (yellow label).
(2) At least three 14-day intervals identified a dose-dense anthracycline regimen: 14-day delays followed by (i) several 21-day intervals were tagged as anthracycline-docetaxel regimens (orange label); (ii) several seven-day intervals were tagged as anthracycline-paclitaxel regimens (red label).
(3) Any 21-day intervals followed by seven-day intervals were inferred to correspond to anthracycline-paclitaxel regimens (red label).

(4) The remaining 21-day intervals could reflect treatment with either anthracycline or docetaxel. Before March 2012, docetaxel was identifiable in the SNDS because it was part of a special reimbursement process in France, called "*list en sus*", for costly and innovative molecules. A 21-day interval before March 2012 not reimbursed as docetaxel was tagged as an anthracycline regimen. Hence, sequences of 21-day intervals before March 2012 were fully identifiable as anthracyclines-docetaxel (orange label), anthracyclines (dark blue label) or docetaxel (light blue label). After March 2012, it was no longer possible to distinguish between anthracyclines and docetaxel: sequences of 21-day intervals were, therefore, tagged as "Unknown" (gray label).

(5) Finally, combinations of 7, 14 and 21-day intervals between CT sessions not classified by the above rules were classified as "other" (green label).

The number of cycles of CT was calculated as the number of anthracycline or docetaxel sessions plus a third of the number of paclitaxel sessions (rounded downwards), and is presented in the third column of Fig. S2.

**Endocrine therapy (ET)**

We separated ET for cancer treatment from ET for fertility preservation procedures (FPP), by excluding (i) GnRH agonist deliveries during CT (assumed to be prescribed for ovarian protection), (ii) tamoxifen and AI deliveries in the six weeks preceding any embryo or oocyte cryopreservation (assumed to be prescribed for hormonal stimulation, Table S5) and (iii) all GnRH agonist deliveries for patients with less than three deliveries of GnRH agonists in total. A patient was considered to be treated with ET if she had at least one ET delivery between 250 days before to up to 365 days after her BC index surgery, in accordance with clinical practices.

*ET start date* was defined as the date of the first ET delivery within these dates, and the patient was considered to have been treated with neoadjuvant ET (NET) if ET start date was before the BC index surgery; adjuvant otherwise. We prevented early prescriptions of ET from being considered as NET, by not tagging patients whose NET start date was less than two months from index surgery as receiving NET; and their ET start date was postponed to the date of the first ET delivery after index surgery. No *ET end date* was defined, because ET is a long-term treatment (five to years) and we did not have access to seven years of outpatient care history for all the patients.

In cases of delivery of any ET molecule approved for use in the metastatic setting, such as toremifene, fulvestrant, or formestane (TableS2), the patient was assumed to have distant metastases, and all subsequent ET deliveries were excluded from calculation of the ET regimen.

**Targeted therapy (TT)**

The *TT start date* was defined as the date of the first session of anti-*HER2* treatment within the dates considered; and patients were considered to have been treated with neoadjuvant targeted therapy (NTT) if the TT start date was before the BC index surgery date; adjuvant otherwise. *Anti-HER2 end date* was defined as the date of the last session before a discontinuation of sessions for at least six months.

In cases of delivery of any TT molecule approved for use in the metastatic setting, such as lapatinib, bevacizumab, everolimus, palbociclib, or alpelisib (Table S2), the patient was assumed to have distant metastases, and *anti-HER2 end date* was considered to be the last date of anti-*HER2* TT before the delivery of these drugs.

Intervals between consecutive TT sessions were rounded to either 7 or 21 days. The *number of cycles of anti-HER2 TT* was calculated as the number of 21-day intervals between sessions, plus a third of the number of sessions separated by intervals of seven days, rounded down.

The diagnostic code does not distinguish between sessions of TT combined with chemotherapy and sessions of TT alone. We inferred that the patients treated with TT also received chemotherapy unless their ET start date was before the second session of targeted therapy, in which case they were said to be treated with "targeted therapy – endocrine therapy (no chemotherapy)". In accordance with standard clinical practices, patients treated with neoadjuvant targeted therapy (NTT) were not assumed to have received adjuvant CT, unless there was evidence of CT use in an adjuvant setting (CT-only sessions or reporting of the use of CT molecules from the "list en sus"). Chemotherapy regimens, the number of cycles of CT, and targeted therapy + chemotherapy regimens were inferred with a specific algorithm detailed in Fig. S3.

CT/TT sessions were classified as chemotherapy-only sessions (CT sessions without TT; gray pictogram) and targeted therapy +/- chemotherapy sessions (denoted as TT+/-CT; orange pictogram), defined as TT sessions that may or may not have included CT. Hospital stays (day hospital or long-term hospitalizations) containing CT/TT sessions were characterized by their start date, end date and the number of sessions. Patients with any hospital stay for which the number of sessions was inconsistent (*i.e.* more than one session per day) were excluded from the cohort. We then distinguished two categories of patients:

- Patients whose CT/TT sessions all took place at the day hospital, such that the exact date of each CT/TT session was known. This category included most of the patients. Intervals between consecutive CT/TT sessions were calculated as the difference in CT/TT session dates, rounded to 1, 7, 14 or 21 days.
- Patients for whom at least one CT/TT session date was unknown, making it impossible to calculate all the intervals between consecutive 2 CT/TT sessions directly. This category included a minority of patients. We inferred the sequence of CT/TT intervals for these patients from the most frequent sequence of intervals for patients in the first category matching the available data concerning intervals between CT/TT sessions for the patient. If the inferred sequence of intervals was not among the 50 most frequent sequences of intervals for patients in the first category; the inference was tagged as uncertain; and the patient was excluded from the cohort.

Daily chemotherapy sessions (defined as a one-day interval between consecutive sessions) were tagged as indicating a suspicion of metastatic disease. If at least three consecutive daily CT sessions were identified within six months of BC index surgery, the patient was considered to have had stage IV BC at diagnosis and was excluded from the cohort. If at least three consecutive daily CT sessions were identified six months or more after BC index surgery, the patient was assumed to have distant metastases or disease progression, and all subsequent CT sessions were excluded from the calculation of CT regimen.

TT + chemotherapy regimens were then inferred from the sequence of intervals between CT/TT sessions, and the type of session (CT only or TT+/-CT), as detailed in the second column of the Fig. S3. Chemotherapy regimens were directly inferred from TT + chemotherapy regimens (third column of Fig. S3).

The number of cycles of CT was inferred from the number of 7-day, 14-day and 21-day intervals between sessions, as detailed in the fourth column of Fig. S3. This number may not be known, in some cases, due to the impossibility of distinguishing between TT-only and TT+CT sessions. The number of cycles of TT could always be calculated, as the number of 21-day intervals between TT+/-CT sessions plus a third of the number of seven-day intervals between TT+/-CT sessions (rounded down), as detailed in the fifth column of Fig. S3.
