## Supplemental Figure S3 for "The French Early Breast Cancer Cohort (FRESH): a resource for breast cancer research and evaluations of oncology practices based on the French National Healthcare System Database (SNDS)"

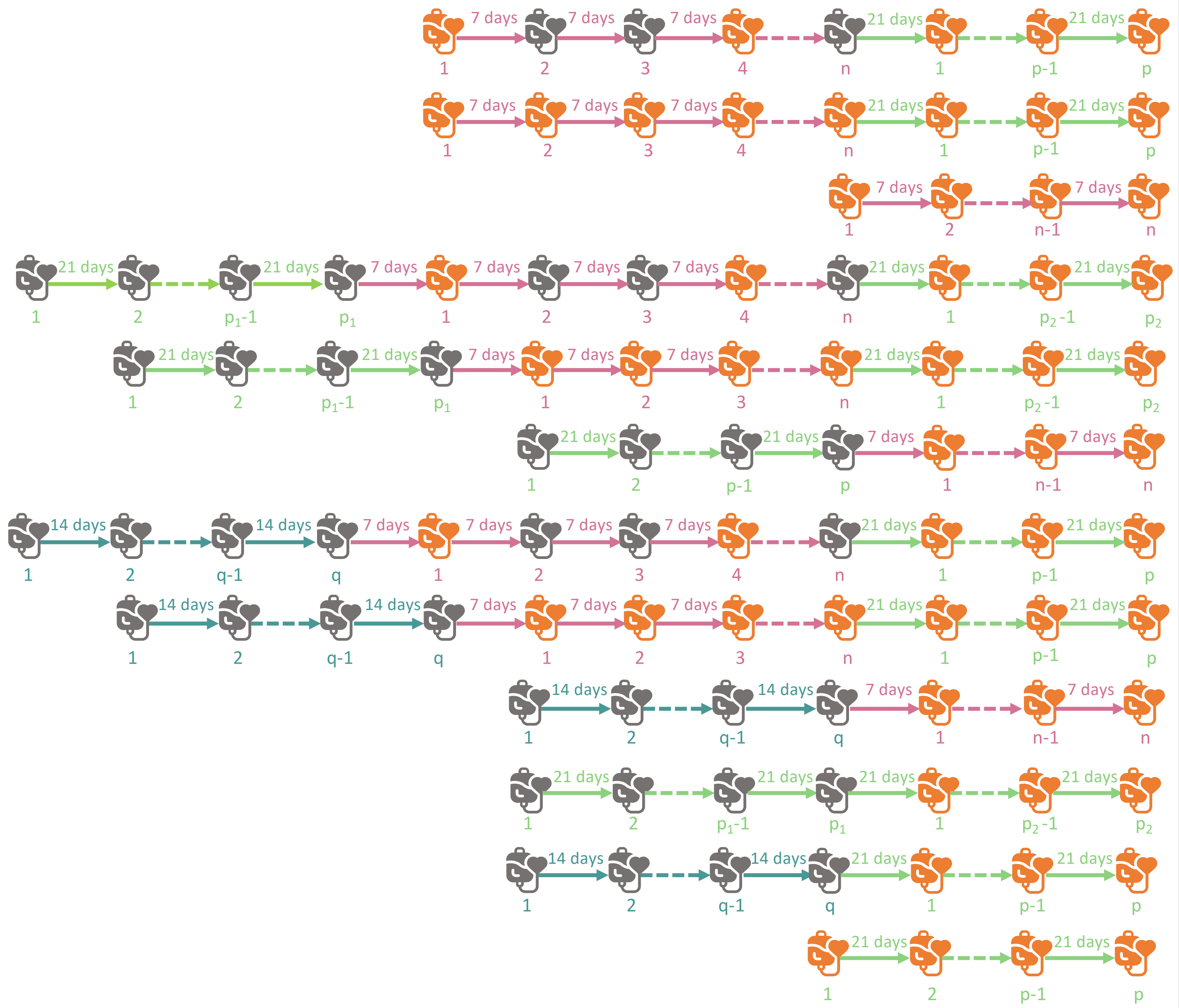

| Sequence of intervals | Targeted therapy + Chemotherapy regimen | Chemotherapy regimen | Number of cycles of chemotherapy | Number of cycles of targeted therapy |
|---|---|---|---|---|
| | paclitaxel-TT (Tolaney) | paclitaxel | E(n/3) | E(n/3) + p |
| | paclitaxel-TT (Tolaney) | paclitaxel | E(n/3) | E(n/3) + p |
| | paclitaxel-TT (Tolaney) | paclitaxel | Unknown | E(n/3) |
| | anthracyclines/paclitaxel-TT | anthracyclines/paclitaxel | $p_1$ + E(n/3) | E(n/3) + $p_2$ |
| | anthracyclines/paclitaxel-TT | anthracyclines/paclitaxel | $p_1$ + E(n/3) | E(n/3) + $p_2$ |
| | anthracyclines/paclitaxel-TT | anthracyclines/paclitaxel | Unknown | E(n/3) + p |
| | anthracyclines/docetaxel-TT | anthracyclines/docetaxel | q + E(n/3) | E(n/3) + p |
| | anthracyclines/docetaxel-TT | anthracyclines/docetaxel | q + E(n/3) | E(n/3) + p |
| | anthracyclines/docetaxel-TT | anthracyclines/docetaxel | Unknown | E(n/3) |
| | anthracyclines/docetaxel-TT | anthracyclines/docetaxel | Unknown | $p_2$ |
| | anthracyclines/docetaxel-TT | anthracyclines/docetaxel | Unknown | p |
| | docetaxel-TT | docetaxel | Unknown | p |
| Any other | other | other | Unknown | E(n/3) + p where n is the number of 7-day interval Targeted therapy +/- chemotherapy session and p is the number of 21-day interval Targeted therapy +/- chemotherapy session |

**Notations :**

- Chemotherapy only session
- Targeted therapy +/- chemotherapy session
- Rounded interval
- n Number of 7-day interval sessions
- q Number of 14-day delay sessions
- p Number of 21-day interval sessions
