## Supplemental Table S1 for "The French Early Breast Cancer Cohort (FRESH): a resource for breast cancer research and evaluations of oncology practices based on the French National Healthcare System Database (SNDS)"

Supp table 1 BC

| **Medical data** | **ICD-10** |
|---|---|
| Carcinoma *in situ* of the breast | D05 |
| Lobular carcinoma *in situ* of the breast | D050 |
| Intraductal carcinoma *in situ* of the breast | D051 |
| Other carcinoma *in situ* of the breast | D057 |
| Unspecified type of carcinoma *in situ* of the breast | D059 |
| Malignant breast tumor | C50 |
| Malignant neoplasm of nipple and areola | C500 |
| Malignant neoplasm of central portion of the breast | C501 |
| Malignant neoplasm of upper-inner quadrant of the breast | C502 |
| Malignant neoplasm of lower-inner quadrant of the breast | C503 |
| Malignant neoplasm of upper-outer quadrant of the breast | C504 |
| Malignant neoplasm of lower-outer quadrant of the breast | C505 |
| Malignant neoplasm of axillary tail of the breast | C506 |
| Malignant neoplasm of overlapping sites of the breast | C508 |
| Malignant neoplasm of the breast of unspecified site | C509 |

**Table S1** : ICD-10 diagnosis codes used to identify breast cancer
Abbreviations : ICD 10 = International Statistical Classification and Related Health Problems – $10^{th}$ revision.
