## Supplemental Table S2 for "The French Early Breast Cancer Cohort (FRESH): a resource for breast cancer research and evaluations of oncology practices based on the French National Healthcare System Database (SNDS)"

| | Medical Data | ICD-10 | Medical Data | ICD-10 |
|---|---|---|---|---|
| **Diagnosis code of other cancer** | Lip | C00 | Ureter, bladder, other unspecified urinary organs | C66, C67, C68 |
| | Tongue | C01, C02 | Eye and adnexa, meninges | C69, C70 |
| | Gum | C03 | Brain | C71 |
| | Floor of mouth | C04 | Spinal cord, cranial nerves and other parts of CNS | C72 |
| | Palate, other and unspecified parts of the mouth | C05, C06 | Thyroid gland | C73 |
| | Parotid and other salivary glands | C07, C08 | Adrenal gland, endo glands and related structures, other ill-defined sites | C74, C75, C76 |
| | Tonsil | C09 | | |
| | Oro/naso/hypopharynx | C10, C11, C13 | Without specification of site | C80 |
| | Pyriform sinus | C12 | Hodgkin lymphoma | C81 |
| | Lip, oral cavity and pharynx | C14 | Follicular lymphoma | C82 |
| | Esophagus, stomach | C15, C16 | Non-follicular lymphoma | C83 |
| | Small intestine, colon | C17, C18 | Mature T/NK-cell lymphomas | C84 |
| | Rectosigmoid junction, rectum | C19, C20 | Other and unspecified types of non-Hodgkin lymphoma | C85 |
| | Anus and anal canal | C21 | Malig immunoproliferative dis and certain oth B-cell lymph | C88 |
| | Liver and intrahepatic bile ducts | C22 | Multiple myeloma and malignant plasma cell neoplasms | C90 |
| | Gallbladder, other and unsp. parts of biliary tract | C23, C24 | Lymphoid leukemia | C91 |
| | Pancreas, other and ill-defined digestive organs | C25, C26 | Myeloid leukemia | C92 |
| | Nasal cavity and middle ear, accessory sinuses | C30, C31 | Monocytic leukemia | C93 |
| | Larynx, trachea, bronchus | C32, C33, C34 | Other leukemias of specified cell type | C94 |
| | Thymus, heart, mediastinum and pleura | C37, C38 | Leukemia of unspecified cell type | C95 |
| | Respiratory system and intrathoracic organs | C39 | Other & unspecified malignant neoplasm of lymphoid, hematopoietic cells and tissues | C96 |
| | Bone and articular cartilage of limbs or unsp. sites | C40, C41 | | |
| | Mesothelioma, Kaposi's sarcoma | C45, C46 | Carcinoma *in situ* of oral, digestive, ear, respiratory, genital, unspecified organs | D00, D01, D02, D07, D09 |
| | Peripheral nerves and autonomic nervous system | C47 | | |
| | Retroperitoneum and peritoneum | C48 | Neoplasm of uncertain behavior of oral cavity and digestive organs | D37, D38, D39, D40, D41, D42, D43, D44 |
| | Other connective and soft tissue | C49 | | |
| | Vulva, vagina | C51, C52 | | |
| | Corpus uteri, part unspecified | C54, C55 | Myelodysplastic syndromes | D46 |
| | Ovary, other and unsp. female genital organs | C56, C57 | Other neoplasm of uncertain behavior of lymphoid, hematopoietic cells and tissues | D47 |
| | Placenta | C58 | | |
| | Penis, other and unsp. male genital organs | C60, C63 | Neoplasm of uncertain behavior of other and unsp sites | D48 |
| | Kidney, renal pelvis | C64, C65 | | |

| | Medical Data | ATC | Medical Data | ATC |
|---|---|---|---|---|
| **Chemotherapy or immunotherapy molecules not indicated for breast cancer** | BENDAMUSTINE | L01AA09 | RITUXIMAB | L01XC02 |
| | BUSULFAN | L01AB01 | CETUXIMAB | L01XC06 |
| | CARMUSTINE | L01AD01 | PANITUMUMAB | L01XC08 |
| | FOTEMUSTINE | L01AD05 | IPILIMUMAB | L01XC11 |
| | RALTITREXED | L01BA03 | TEMSIROLIMUS | L01XE09 |
| | PEMETREXED | L01BA04 | TOPOTECAN | L01XX17 |
| | CLADRIBINE | L01BB04 | IRINOTECAN | L01XX19 |
| | CLOFARABINE | L01BB06 | ARSENIC TRIOXIDE | L01XX27 |
| | CYTARABINE | L01BC01 | BORTEZOMIB | L01XX32 |
| | AZACITIDINE | L01BC07 | ALEMTUZUMAB | L04AA34 |
| | TRABECTEDIN | L01CX01 | LENALIDOMIDE | L04AX04 |
| | IDARUBICIN | L01DB06 | | |

**Table S2** : ICD-10 and ATC codes used to identify cancer at another site.

Abbreviations : ICD-10 = International Statistical Classification and Related Health Problems – 10th revision ; ATC = Anatomical Therapeutic and Chemical
