## Supplemental Table S3 for "The French Early Breast Cancer Cohort (FRESH): a resource for breast cancer research and evaluations of oncology practices based on the French National Healthcare System Database (SNDS)"

Supp_table_3_Meta

| | | Medical data | ICD-10 | ATC |
|---|---|---|---|---|
| **Diagnosis code for metastatic disease** | | Metastatic disease | C770, C771, C772, C774, C775, C778, C779, C78, C780, C781, C782, C783, C784, C785, C786, C787, C788, C79, C790, C791, C792, C793, C794, C795, C796, C797, C798, C799 | |
| **Molecules indicated only for metastatic disease** | **Chemotherapy** | CAPECITABINE | | L01BC06 |
| | | CARBOPLATIN | | L01XA02 |
| | | CISPLATIN | | L01XA01 |
| | | DOXORUBICIN (only CAELYX®) | | L01DB01 |
| | | ERIBULIN | | L01XX41 |
| | | ETOPOSIDE | | L01CB01 |
| | | GEMCITABINE | | L01BC05 |
| | | MELPHALAN | | L01AA03 |
| | | METHOTREXATE | | L01BA01 |
| | | MILTEFOSINE | | L01XX09 |
| | | MITOMYCIN | | L01DC03 |
| | | MITOXANTRONE | | L01DB07 |
| | | PIRARUBICIN | | L01DB08 |
| | | THIOTEPA | | L01AC01 |
| | | VINBLASTINE | | L01CA01 |
| | | VINCRISTINE | | L01CA02 |
| | | VINDESINE | | L01CA03 |
| | | VINORELBINE | | L01CA04 |
| | ***HER2*- Targeted Therapy** | LAPATINIB | | L01XE07 |
| | **Non-*HER2* Targeted Therapy** | BEVACIZUMAB | | L01XC07 |
| | | BYL719 (ALPELISIB) | | L01XE |
| | | EVEROLIMUS | | L01XE10 |
| | | PALBOCICLIB | | L01XE33 |
| | **Endocrine Therapy** | TOREMIFENE | | L02BA02 |
| | | FORMESTANE | | L02BG02 |
| | | FULVESTRANT | | L02BA03 |
| | **Other** | DEXRAZOXANE | | V03AF02 |
| | | SAMARIUM (153SM) LEXIDRONAM | | V10BX02 |

**Table S3**: ICD-10 and ATC codes used to identify *de novo* metastatic breast cancer.

Abbreviations: ICD 10 = International Statistical Classification and Related Health Problems – 10th revision ; ATC = Anatomical Therapeutic and Chemical Classification.
