## Supplemental Table S4 for "The French Early Breast Cancer Cohort (FRESH): a resource for breast cancer research and evaluations of oncology practices based on the French National Healthcare System Database (SNDS)"

| | | Medical Data | CCAM | ICD-10 | ATC |
|---|---|---|---|---|---|
| **Surgery** | | Mastectomy with axillary surgery | QEFA003, QEF005, QEFA010, QEFA020 | | |
| | | Mastectomy without axillary surgery | QEFA007, QEFA012, QEFA013, QEFA015, QEFA019 | | |
| | | Partial mastectomy with axillary surgery | QEFA001, QEFA008 | | |
| | | Partial mastectomy without axillary surgery | QEFA004, QEFA016, QEFA017, QEFA018 | | |
| | | Axillary surgery without breast surgery | FCFA021, FCFA029 | | |
| **Radiotherapy** | | Appointment for antineoplastic radiation therapy | | Z5100 | |
| | | Radiotherapy preparation session | | Z5101 | |
| | | Radiotherapy session | | Z510 | |
| | | Radiotherapy procedures | ZZNA002, ZZNL001, ZZNL002, ZZNL003, ZZNL004, ZZNL005, ZZNL006, ZZNL009, ZZNL011, ZZNL012, ZZNL013, ZZNL014, ZZNL015, ZZNL016, ZZNL017, ZZNL018, ZZNL019, ZZNL02, ZZNL030, ZZNL031, ZZNL036, ZZNL037, ZZNL039, ZZNL040, ZZNL042, ZZNL043, ZZNL045, ZZNL046, ZZNL048, ZZNL049, ZZNL050, ZZNL051, ZZNL052, ZZNL053, ZZNL054, ZZNL055, ZZNL058, ZZNL059, ZZNL060, ZZNL061, ZZNL062, ZZNL063, ZZNL064, ZZNL065, ZZNL900, ZZNL902, ZZNL903, ZZNL904, ZZNL905, ZZNL906, YYYY016, YYYY021, YYYY023, YYYY045, YYYY046, YYYY047, YYYY048, YYYY049, YYYY050, YYYY051, YYYY052, YYYY053, YYYY054, YYYY055, YYYY056, YYYY080, YYYY081, YYYY099, YYYY101, YYYY109, YYYY122, YYYY128, YYYY136, YYYY141, YYYY151, YYYY152, YYYY166, YYYY175, YYYY197, YYYY1211, YYYY223, YYYY225, YYYY244, YYYY256, YYYY267, YYYY299, YYYY301, YYYY302, YYYY303, YYYY304, YYYY305, YYYY306, YYYY307, YYYY310, YYYY312, YYYY313, YYYY314, YYYY315, YYYY316, YYYY320, YYYY323, YYYY324, YYYY325, YYYY326, YYYY327, YYYY331, YYYY334, YYYY335, YYYY336, YYYY337, YYYY338, YYYY343, YYYY345, YYYY346, YYYY347, YYYY348, YYYY349, YYYY356, YYYY357, YYYY358, YYYY359, YYYY360, YYYY365, YYYY367, YYYY368, YYYY369, YYYY370, YYYY371, YYYY377, YYYY379, YYYY380, YYYY381, YYYY382, YYYY383, YYYY387, YYYY390, YYYY391, YYYY392, YYYY393, YYYY398, YYYY450, YYYY451, YYYY457, YYYY458, YYYY459, YYYY460, YYYY468, YYYY469, YYYY470, YYYY471, YYYY479, YYYY480, YYYY481, YYYY491, YYYY492, YYYY493, YYYY497, YYYY500, YYYY511, YYYY520, YYYY522, YYYY533, YYYY544, YYYY555, YYYY566, YYYY577, YYYY588, YYYY599 | | |
| **Chemotherapy** | | Encounter for antineoplastic chemotherapy and immunotherapy | | Z511, Z512 | |
| | | Anticancerous agent administration | ZZLF900 | | |
| | | Port implanting | EBLA001 | | |
| | | Cerebral anticancerous agent administration | ABLB006 | | |
| | | Intrathecal anticancerous agent administration | AFLB003, AFLB013 | | |
| | | Upper limb arterial anticancerous agent administration | ECLF005, ECLF006 | | |
| | | Lower limb arterial anticancerous agent administration | EELF004, EELF005 | | |
| | | Liver anticancerous agent *in situ* administration | EDLF014, EDLF015, EDLF016, EDLF017 | | |
| | | Kidney anticancerous agent *in situ* administration | EDLF018, EDLF019, EDLF020, EDLF021 | | |
| | | Cervical/cephalic anticancerous agent administration | EBLF002, EBLF003 | | |
| | | Intrapleural anticancerous agent administration | GGLB001, GGLB008 | | |
| | | Intraperitoneal anticancerous agent administration | HPLB002, HPLB003, HPLB007 | | |
| | | Locoregional anticancerous agent administration with extracorporeal circulation | ZZLF004 | | |
| | | Locoregional anticancerous agent administration without extracorporeal circulation | ZZLF900 | | |
| | | CYCLOPHOSPHAMIDE | | | L01AA01 |
| | | DOCETAXEL (identifiable until March 2012) | | | L01CD02 |
| | | EPIRUBICIN | | | L01DB03 |
| | | FLUOROURACIL | | | L01BC02 |
| | | PACLITAXEL | | | L01CD01 |
| **Targeted Therapy** | ***Anti HER2*** | PERTUZUMAB | | | L01XC13 |
| | | TRASTUZUMAB | | | L01XC03, L01XC14 |
| **Endocrine Therapy** | **Tamoxifen** | TAMOXIFEN | | | L02BA01 |
| | **Aromatase Inhibitor** | ANASTROZOLE | | | L02BG03 |
| | | EXEMESTANE | | | L02BG06 |
| | | LETROZOLE | | | L02BG04 |
| | **GnRH agonists** | GOSERELIN | | | L02AE03 |
| | | LEUPRORELIN | | | L02AE02 |
| | | TRIPTORELIN | | | L02AE04 |

**Table S4**: CCAM, ICD-10 and ATC codes used to identify breast cancer treatments.

Abbreviations: CCAM = French Common Classification of Medical Procedures; ICD 10 = International Statistical Classification and Related Health Problems – 10th revision; ATC = Anatomical Therapeutic and Chemical Classification; HER2=human epidermal growth factor receptor 2.
