## Supplemental Table S5 for "The French Early Breast Cancer Cohort (FRESH): a resource for breast cancer research and evaluations of oncology practices based on the French National Healthcare System Database (SNDS)"

Supp_table_5_FPP

| Medical data | CCAM | ATC | CIP | UCD |
|---|---|---|---|---|
| Celioscopic oocyte extraction | JJFC011 | | | |
| Vaginal oocyte extraction | JJFJ001 | | | |
| Chorionic gonadotrophin | | G03GA01 | | |
| Choriogonadotropin alfa | | G03GA08 | | |
| Short-acting gonadotrophin agonists | | | 3285026, 3285032, 3287924, 3285061, 3285055, 3328563, 3535210 | 9109419, 9112077, 9112060, 9148603, 9219958 |
| Gonadotrophin antagonist | | | 3717790, 3517815, 3553018, 3553024 | 9218812, 9218829, 9233591 |

**Table S5**: CCAM, ATC, CIP and UCD codes used to identify embryo or oocyte cryopreservation

Abbreviations: CCAM = French Common Classification of Medical Procedures; ATC = Anatomical Therapeutic and Chemical Classification; CIP (*Code Identifiant de Présentation*); UCD (*Unités Communes de Dispensation*)
