## Supplemental Table S6 for "The French Early Breast Cancer Cohort (FRESH): a resource for breast cancer research and evaluations of oncology practices based on the French National Healthcare System Database (SNDS)"

Supp_table_6_Diag

| Medical data | | CCAM |
|---|---|---|
| **Breast core biopsy** | | QEHA001, QEHA002, QEHB002, QEHJ001, QEHH001, QEHJ005, QEHJ006, QEHH002, QEHH015, QEHJ004 |
| **Fine needle aspiration cytology** | | QEHB001, QEHJ002, QEHJ003, QEHH003, QEHP002 |
| **Breast imaging procedures** | Mammography | QEQK001, QEQK004, QEQK005 |
| | Mammary ultrasound | QEQM001 |
| | Mammary MRI | QEQJ001, QEQN001 |
| | CT scan | QEQH002, QEQK006 |
| | Galactography | QEQH001 |

**Table S6: CCAM codes used to identify diagnostic procedures for breast cancer**

Abbreviations: CCAM = French Common Classification of Medical Procedures; CT = computed tomography.
